# DiscoDivas: Leveraging genetic ancestry continuum information to interpolate PRS for admixed populations

**DOI:** 10.1101/2024.11.09.24316996

**Authors:** Yunfeng Ruan, Rohan Bhukar, Aniruddh Patel, Satoshi Koyama, Leland Hull, Buu Truong, So Mi Cho, Whitney Hornsby, Haoyu Zhang, Nilanjan Chatterjee, Pradeep Natarajan

## Abstract

The relatively low representation of admixed populations in both discovery and fine-tuning individual-level datasets limits polygenic risk score (PRS) development and equitable clinical translation for admixed populations. Under the assumption that the most informative PRS model for a genetically homogeneous sample varies linearly in an ancestry continuum space, we introduce a Genetic **Dis**tance-assisted PRS **Co**mbination Pipeline for **Div**erse Genetic **A**ncestrie**s** (**DiscoDivas**) to interpolate a harmonized PRS for diverse, especially admixed, genetic ancestries, leveraging multiple PRS models fine-tuned within existing samples, which are mostly of single ancestry, and genetic distance. DiscoDivas treats genetic ancestry as a continuous variable and does not require shifting between different models when calculating PRS for different ancestries. We generated PRS with DiscoDivas and the current conventional method, i.e. fine-tuning multiple GWAS PRS using the matched or similar genetic ancestry samples. DiscoDivas generated a harmonized PRS of the accuracy comparable to or higher than the conventional approach, with the greatest advantage exhibited in admixed individuals.

## Introduction

Individuals who are not of European ancestries remain underrepresented in genome-wide association studies (GWAS), which at least partly explains why polygenic risk score (PRS) performance is generally reduced in this population when compared with individuals of European ancestries^1^. Within the constraints of existing data, the current principal solution to increase the PRS accuracy among non-European individuals is to fine-tune a combination of PRS derived from multiple populations or multiple traits with the individual-level data of a training cohort^2–6^. However, PRS accuracy decays as the genetic distance between the fine-tuning and testing samples increases^7^. Relative to the vast diversity across the genetic ancestry continuum, the existing and near-term individual-level datasets that can be used for fine-tuning PRS combinations remains very sparse. Most existing individual-level genotype data are mainly collected from single-ancestry populations and therefore admixed populations are left underrepresented or are largely excluded from analysis ^8–11^. Additionally, fine-tuning and testing samples that are labeled as “from the same superpopulation” are often truly genetically heterogeneous ^10,12–15^, leading to variable accuracy within such samples.

PRS analysis across diverse ancestries may also be limited by inconsistency. The raw PRS distributions of the same model varies by ancestry and therefore the raw PRS values for individuals of different genetic ancestries should not be directly compared without ancestry correction^16–18^. Although prior research^16,18,19^ has shown that regressing out the top principal components (PCs) of ancestry from the PRS can unify the PRS distributions of different ancestries (i.e., the mean and standard deviation of corrected PRS sampled from different populations can become very close), the accuracy of one PRS model still varies across different genetic ancestries. In the application of PRS across diverse ancestries, one would have to use one PRS model for all the individuals, causing inconsistent PRS accuracy, or use several discrete PRS models for different individuals approximating superpopulations also causing inconsistent PRS modelling and accuracy.

Given these issues and the increasing clinical use of PRS^20–22^, PRS generation for diverse and admixed genetic ancestries with more consistent accuracy and more unified PRS distributions is critically needed. Several methods have been developed to incorporate local ancestry inference information into PRS models to increase prediction accuracy in admixed populations^23,24^. However, the effectiveness of these approaches is constrained by the lack of widely accepted standards for local ancestry analysis, accuracy of local ancestry inference, and by their reliance on reference panels^25,26^. Moreover, methods for fine-tuning PRS models have advanced rapidly in recent years, including utilizing summary-level genetic data^27^. The advances in PRS methods and the on-going effort to collect genetic data from diverse populations allow PRS to be trained in cohorts with ancestry profiles that differ from those of the reference panels, presenting challenges for reference-dependent methods to fully utilize these models. Finally, if causal SNPs and their effect sizes can be accurately identified and estimated, neither observed marginal effect sizes across populations nor PRS accuracy decay across populations may pose a challenge in constructing PRS for diverse populations. However, the current genetic data for many traits is insufficient to reach this ideal. Therefore, it remains important to consider population differences and the notion of PRS accuracy decay with genetic distance remains essential when constructing PRS across multiple populations.

In light of the present challenges and opportunities, we devised a method, DiscoDivas, a Genetic ***Dis***tance-assisted PRS ***Co***mbination Pipeline for ***Div***erse Genetic ***A***ncestrie***s***, to generate PRS across the genetic ancestry continuum directly from the fine-tuning cohorts, which eliminates the need to align individuals with external reference panels and further enhances PRS performance beyond what is achieved by existing methods based on a single fine-tuning dataset. This method is based on the recent observations^7,28^ that the PRS accuracy in the testing data decays approximately linearly as the genetic distance between the fine-tuning data and testing data increases, and that the genetic distance can be approximated by Euclidean distance of PC based on the global ancestries^7^. Further support comes from additional findings: accuracy of PRS model trained in European population similarly declines linearly as the proportion of European genetic ancestry decreased in African American testing samples^29^ and that principal component of ancestry has also used to determine genetic ancestry proportion^30^. Together, these observations indicate that PCs can provide meaningful information for modeling and anticipating PRS prediction accuracy, most plausibly via a linear relationship. Based on the above observations, we assumed that the correlation of the most informative PRS models for two samples of different ancestries drops linearly as the Euclidean distance of global ancestry-based PCs between them increases, and therefore, the most informative PRS model for a genetically homogenous sample can be linearly interpolated from the currently available PRS models that are fine-tuned in the ancestries surrounding it in the global ancestry-based PC space with the interpolation coefficients mainly based on the Euclidean distance of the PCs.

In summary, DiscoDivas calculates PRS for diverse and admixed genetic ancestries, whose currently available genetic data may not be sufficiently powerful alone to train a PRS model, by linearly interpolating the multiple PRS fine-tuned in ancestries whose genetic data are more available. We evaluated its performance in simulated and empiric data.

## Results

### Overview of DiscoDivas

DiscoDivas combines PRS fine-tuned in different fine-tuning samples - generally from different single-ancestry populations - to linearly interpolate PRS for individuals of diverse genetic ancestries, treating ancestry as a continuous variable. Based on the recent observations ^7,28–30^, we assumed that the best PRS model, i.e. a SNP effect size vector, for an ancestry representation can be linearly interpolated from other PRS models fine-tuned in other ancestries with the additional consideration of the genetic distance between the samples (Figure 1).

**Figure 1:**
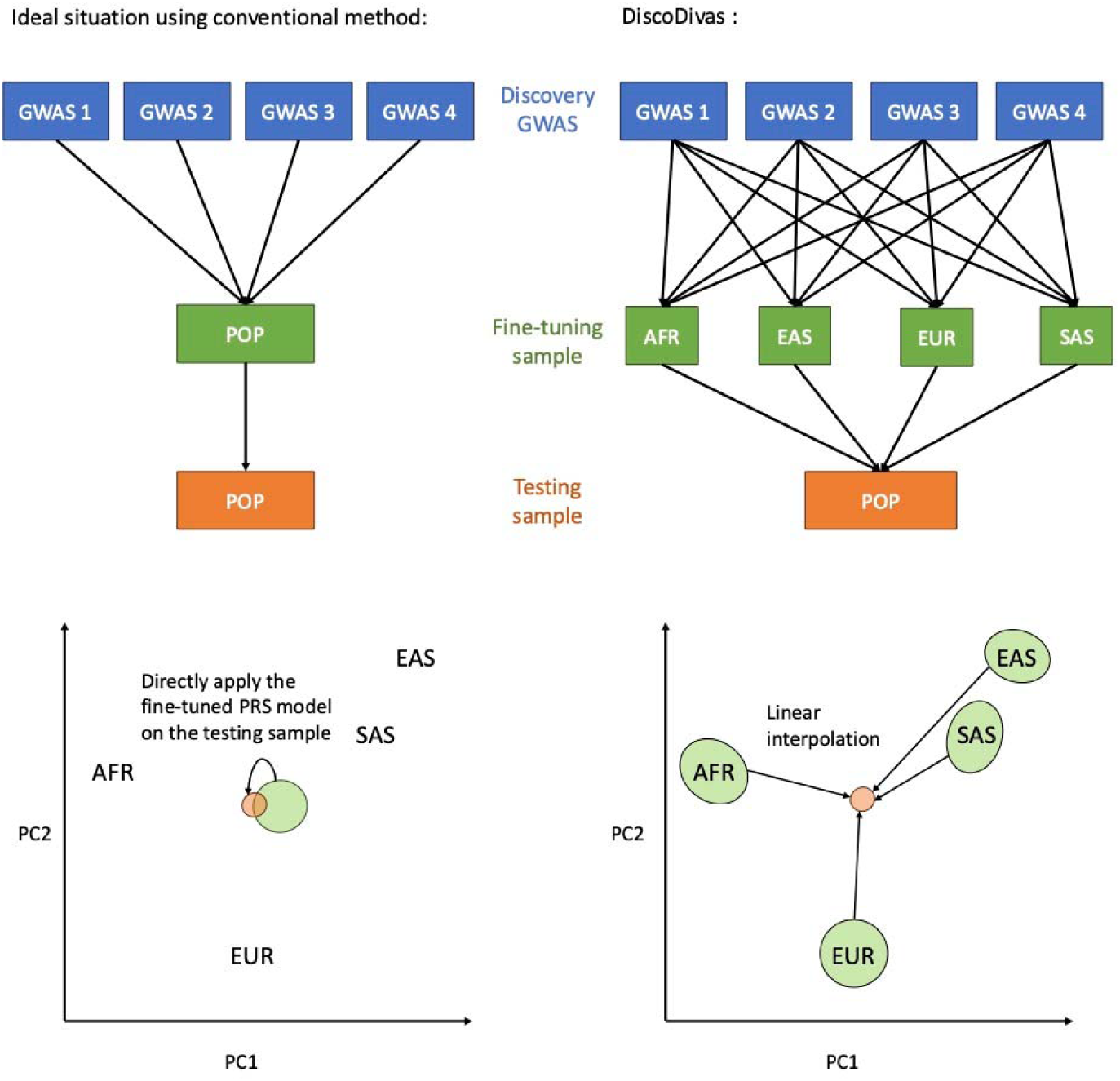
The workflow of comparing DiscoDivas with the existing method. Left: The ideal situation for the existing method is to fine-tune a PRS model that contains multiple GWAS with matched fine-tuning data, which is not currently available for many under-represented populations. Right: DiscoDivas first fine-tunes the PRS in the available ancestries, which are currently AFR, EAS, EUR, and SAS, and interpolates PRS for diverse ancestry groups based on these fine-tuned PRS. In this plot, POP refers to any ancestry for which the PRS is to be calculated. The bottom 2 panels showed the PC distribution of fine-tuning datasets (green circles) and the testing data (orange circles). While more PCs were used in the analysis, only PC1 and PC2 are shown here for clarity.

Under the same principle of interpolating the PRS models, the best PRS for an individual can be interpolated from several PRS calculated using the PRS models fine-tuned in other ancestries for this individual. Since generating individual-specific PRS models in a testing dataset causes redundant calculation and given the difficulty of normalizing information from different datasets, we combine the PRS instead of the SNP models. The PRS of individual *i PRS_i_* in the testing sample is a linear combination of PRS based on the PRS models fine-tuned in different fine-tuning

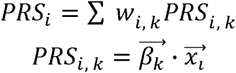

Where 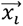 is the genotype vector of the testing individua testing individual *i*; *PRS_i,k_* is the PRS of testing individual *i* calculated using the PRS model 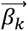 fine-tuned in the fine-tuning *k*; *w_i,k_* PC Euclidean distance between the testing individual ’ and median point of the fine-tuning sample *k*, *D_i,k_*. Note that the input PRS and PCs should be properly harmonized: all the individuals are projected onto the same PC space based on a global ancestry reference panel and the PRS input *PRS_i_*,*_k_* is the raw PRS regressed out the top PCs and then standardized to mean = 0 and sd =1. Additionally, we recommend including all available discovery GWAS when generate the input PRS *PRS_i_*,*_k_* to maximize the PRS accuracy, as indicated in Figure 1. Nevertheless, DiscoDivas is a flexible framework that also accommodates input *PRS_i_*,*_k_* derived from different sets of discovery GWAS and even generated using different PRS methods.

In addition to the PC distances, other factors are included in the model. First, since some fine-tuning samples are more correlated than others (e.g., EAS and SAS are more correlated than AFR and EUR), the combination coefficients should be further modified by these correlations, which can also be extracted from the PC Euclidean distances. Second, since PRS fine-tuned in each of the fine-tuning samples may be of differing qualities (e.g., when the PRS model fine-tuned in different samples are based on GWAS of different sample sizes or populations), the quality of the PRS trained with each of the training data will vary and should be taken into account when combining the PRS. Thus, the combination coefficient *w_i,k_* in the previous formula is a function of multiple factors:

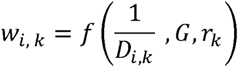

where 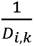 is the reciprocal of PC Euclidean distance between the individual *i* and the fine-tuning sample *k*; *G* is the matrix of PC Euclidean distance between fine-tuning samples; *r_k_* is the parameter describing the quality of training fine-tuning samples. A more detailed description of defining *w_i,k_* is given in the method section entitled ‘Methodological Details of DiscoDivas’.

The PRS input for DiscoDivas in this study was the multi-GWAS PRS fine-tuned in Africa (AFR), East Asian (EAS), European (EUR), and South Asian (SAS) fine-tuning samples with the conventional method pipeline as mentioned in the following section titled “Overview of multi-population GWAS PRS model”). The interpolation from these four PRS is based on the PCA calculated using the 1000 Genomes^31^ reference panel. For most of the PRS analysis conducted in in the present study, the input PRS of DiscoDivas are based on the same set of discovery GWAS and the fine-tuning datasets are sufficiently large to generate a stable result. Therefore, we assumed that all the input PRS can be viewed as of equal quality and their parameter for PRS quality _k_ can be viewed as a constant value in the present study.

### Overview of multi-population GWAS PRS model

A common approach for constructing PRS is to include as much genome-wide association study (GWAS) summary statistic data as possible in the discovery data^5,32,33^. The GWAS data is typically then processed by PRS methods that will adjust the SNP effect size using a set of hyper-parameters. Individual-level data of an independent fine-tuning sample is used to fine-tune the hyper-parameters across PRS methods and the combination of the fine-tuned PRS. The resulting PRS is expected to perform the best in samples of matched ancestry with the fine-tuning sample.

The current approach, as shown in the left panels of Figure 1, is to use the multi-GWAS PRS fine-tuned in the matched sample or the closest approximation when the matched sample is unavailable. The pipeline of adjusting SNP effect sizes and combining information from different GWAS varies widely. Without loss of generality, we built the following pipeline as a representation of the current conventional method: we first adjusted the SNP effect size of each of the summary statistical GWAS datasets by a Bayesian method and then chose the most predictive PRS from all the PRS generated under different hyper-parameters. For simulated GWAS data, we used PRS-CS^34^ to adjust the SNP effect size and LDpred2^35^ for real GWAS. Then we used the fine-tuning data to first select the most predictive PRS based on each GWAS and then to train the linear combination of the most predictive single-GWAS PRS with a linear regression model. The final PRS model generated from each of the fine-tuning datasets is a linear combination of PRS. For the empiric data set, the PRS were fine-tuned controlling for the following covariates: top 20 PCA, sex, and age.

The Individuals from real-life biobanks and simulated individuals based on real-life biobank genotype data were clustered into genetic ancestry Africa (AFR), East Asian (EAS), European (EUR), South Asian (SAS), Admixed American (AMR), Individuals not classified into any of these groups were labeled as Other (OTH). Depending on data availability, either OTH or AMR was used to evaluate PRS performance in admixed populations. In the conventional PRS generation pipeline, fine-tuning datasets were drawn from the major ancestry groups (for single ancestry: AFR, EAS, EUR, SAS; for admixed ancestry: OTH and/or AMR depending on availability). More detailed description of generating PRS models from the one fine-tuning data was given in supplementary method section entitled ‘Methodological Details of PRS Construction Using a Single Fine-tuning Dataset’.

### Simulated data results

Summary-level GWAS used as discovery data were generated based on simulated genotype of AFR, EAS, EUR, and SAS population based on 1000 Genomes^31^ reference as described in the previous publication provided by Zhang et al^6^. Fine-tuning and testing samples were simulated based on UKBB genotype data. From each ancestry group of AFR, EAS, EUR, SAS, and other (OTH) for admixed individuals, 1300 individuals were used as the fine-tuning datasets (See supplementary method section entitled ‘Generating data for simulation analysis’). The phenotype of discovery, fine-tuning, and testing data were generated using the same pipeline and parameters: the phenotypes of 100, 300, 1,000, or 10,000 causal SNPs, a non-genetic factor following a normal distribution across populations and global heritability = 0.6 were simulated. Scenarios of shared causal SNP with effect size constant across different ancestries and shared causal SNP with effect size varying across population are both simulated. We used up to 100,000 simulated individuals from AFR, EAS, EUR, and SAS to generate the discovery summary statistics GWAS dataset with PLINK2^36^ and used 10,000 of the remaining samples as fine-tuning dataset in other downstream analyses.

We primarily focused on the PRS performance in the OTH testing cohort. DiscoDivas, which is based on PRS fine-tuned in AFR, EAS, EUR, and SAS, was compared with the conventional PRS fine-tuned in the matched admixed fine-tuning sample in scenarios of different causal SNP numbers, different discovery GWAS sample sizes, and different causal SNP distribution across ancestry (See Figure 2)

**Figure 2.**
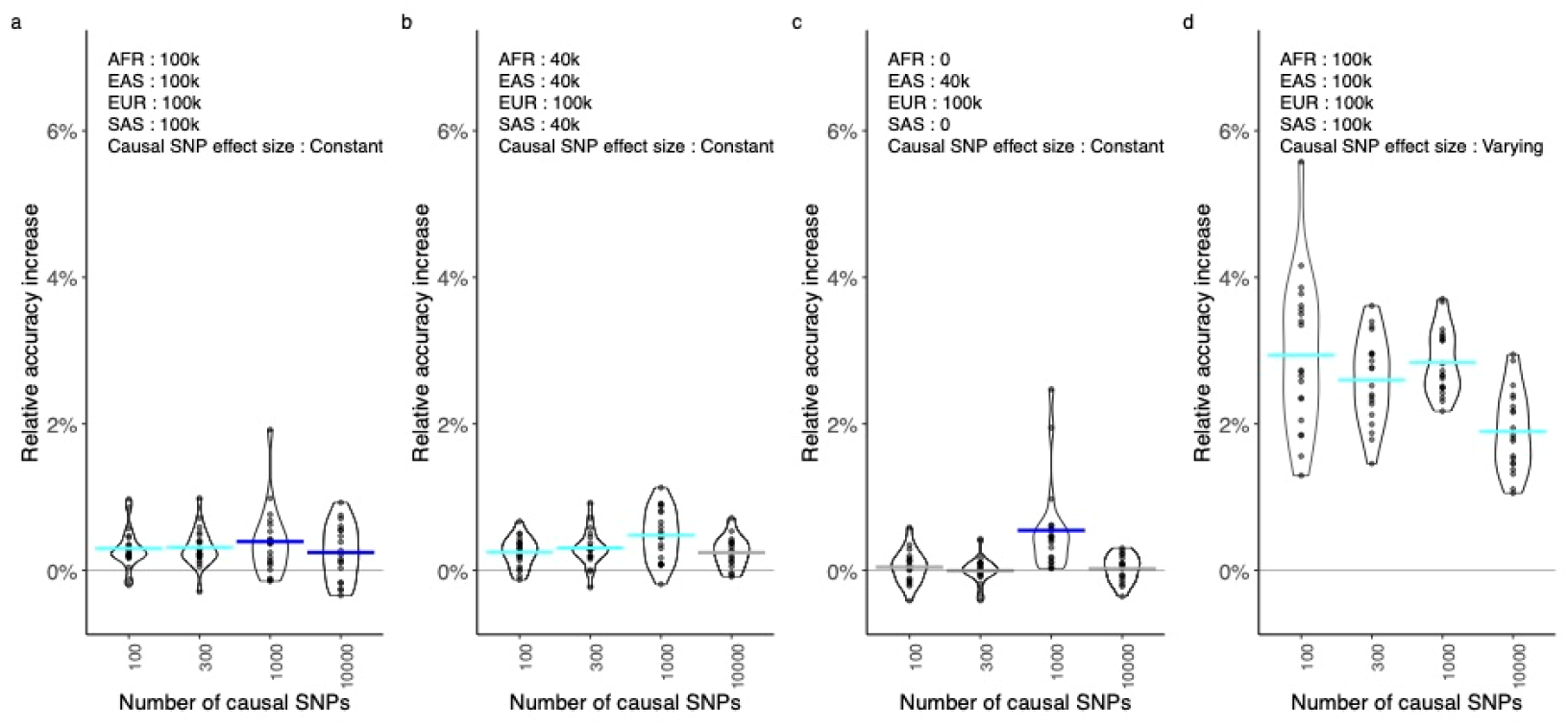
Relative **R**^2^ increase of DiscoDivas over the conventional PRS fine-tuned in a matched sample when tested in admixed individuals. The x-axis shows the simulated number of causal SNPs. The horizontal bar shows the mean relative R^2^ increase and the color of the horizontal bar indicates the *p*-value of the paired t-test of DiscoDivas PRS R^2^ and conventional PRS R^2^, with cyan being *p*-value<0.0005, dark blue being *p*-value<0.05 and grey being *p*-value>0.05. In panels a, b, and c, the causal SNP effect sizes are constant across different populations. The annotation texts on the top of each panel shows the sample size of discovery GWAS of different populations and the distribution of causal SNP effect sizes.

Although the comparison between DiscoDivas and the conventional method of fine-tuning PRS with matched ancestry sample in a single test iteration usually showed no statistical significance due to the small numeric differences, the paired t-test of DiscoDivas R^2^ and the conventional PRS R^2^ over the 20 iterations better clarified significant differences. When effect sizes of causal SNPs were held constant across different ancestries (Figure 2 panel a, b, and c), the PRS generated by DiscoDivas had comparable accuracy with the PRS fine-tuned using matched data. We noticed that when the sample size of non-European discovery GWAS dropped and the dataset was relatively more Eurocentric, the advantage of DiscoDivas became less statistically significant. In Figure 2 panel d, we compared DiscoDivas and the conventional PRS method of fine-tuning the PRS with matched ancestry in the scenario where causal SNPs were shared across all populations, but the effect sizes varied linearly in the PCA space. The advantage of DiscoDivas over conventional PRS method was more obvious in this scenario than when the effect sizes were constant across populations (Figure 2 panel d versus panel a), presumably because personalized PRS combination with DiscoDivas better captured the changing effect sizes for the admixed testing sample. In all the scenarios tested, the advantage of DiscoDivas was least statistically significant when the number of causal SNPs was 10,000 but still significant when the number of causal SNPs was 1,000. Notably, the accuracy of both DiscoDivas and the conventional PRS method was the lowest when the number of causal SNPs was 10,000 (Supplementary Figure 1), indicating that the difference of the two PRS methods became less obvious when the input data became increasingly underpowered.

When predicting the individuals that are usually classified as single ancestries, i.e. AFR, EAS, EUR, and SAS, DiscoDivas showed no statistically significant difference or a slight advantage over the conventional PRS method (Supplementary Figure 2). When predicting AMR individuals, we used admixed fine-tuning data (OTH) to fine-tune the conventional PRS due to the small sample size of the AMR dataset. The PRS performance when testing in the AMR dataset was similar as in admixed data but the statistical significance was weaker, potentially due to the small sample size and the high heterogeneity of the AMR dataset. Besides, DisocDivas showed its clearest advantage over the conventional method of fine-tuning PRS with single dataset when the testing data and the fine-tuning data for the conventional method were of different ancestries (supplementary figure 2). This result underscores the advantage of DiscoDivas in delivering a harmonized and robustly accurate PRS for samples spanning a wide range of genetic ancestries, compared to relying on a single PRS model or switching between multiple discrete PRS models.

### Biobank data results

We downloaded publicly available summary statistical data of body-mass index (BMI), high-density lipoprotein cholesterol (HDL), low-density lipoprotein cholesterol (LDL), total cholesterol (TC), triglycerides (TG), systolic blood pressure (SBP), diastolic blood pressure (DBP), coronary artery disease (CAD), and type 2 diabetes mellitus (DM2) and adjusted the SNP effect size using LDpred2 as described previously^5^.

For the quantitative traits, we used the fine-tuning samples of AFR, EAS, EUR, SAS, and admixed (OTH) ancestry in the UK Biobank (UKBB) to fine-tune the model. The remaining UKBB samples were used as the testing data. The results for empiric quantitative trait data were highly aligned with the simulation results (Figure 3): DiscoDivas showed a robust advantage over the conventional PRS method of fine-tuning PRS with matched or similar ancestry samples when compared across the 7 traits in the OTH testing dataset. When predicting AFR, EAS, EUR, and SAS, DiscoDivas and the conventional PRS method had similar or slightly improved performance. The results of both methods in AMR testing dataset had large deviations due to the small sample size and greater genetic heterogeneity of the AMR data. Consistent with the simulated data, DiscoDivas showed a greater advantage over PRS models fine-tuned on datasets of different ancestry from the testing samples (Supplementary Figure 17). This further supports the potential of DiscoDivas to construct a harmonized and robustly accurate PRS for individuals across a wide range of ancestries.

**Figure 3.**
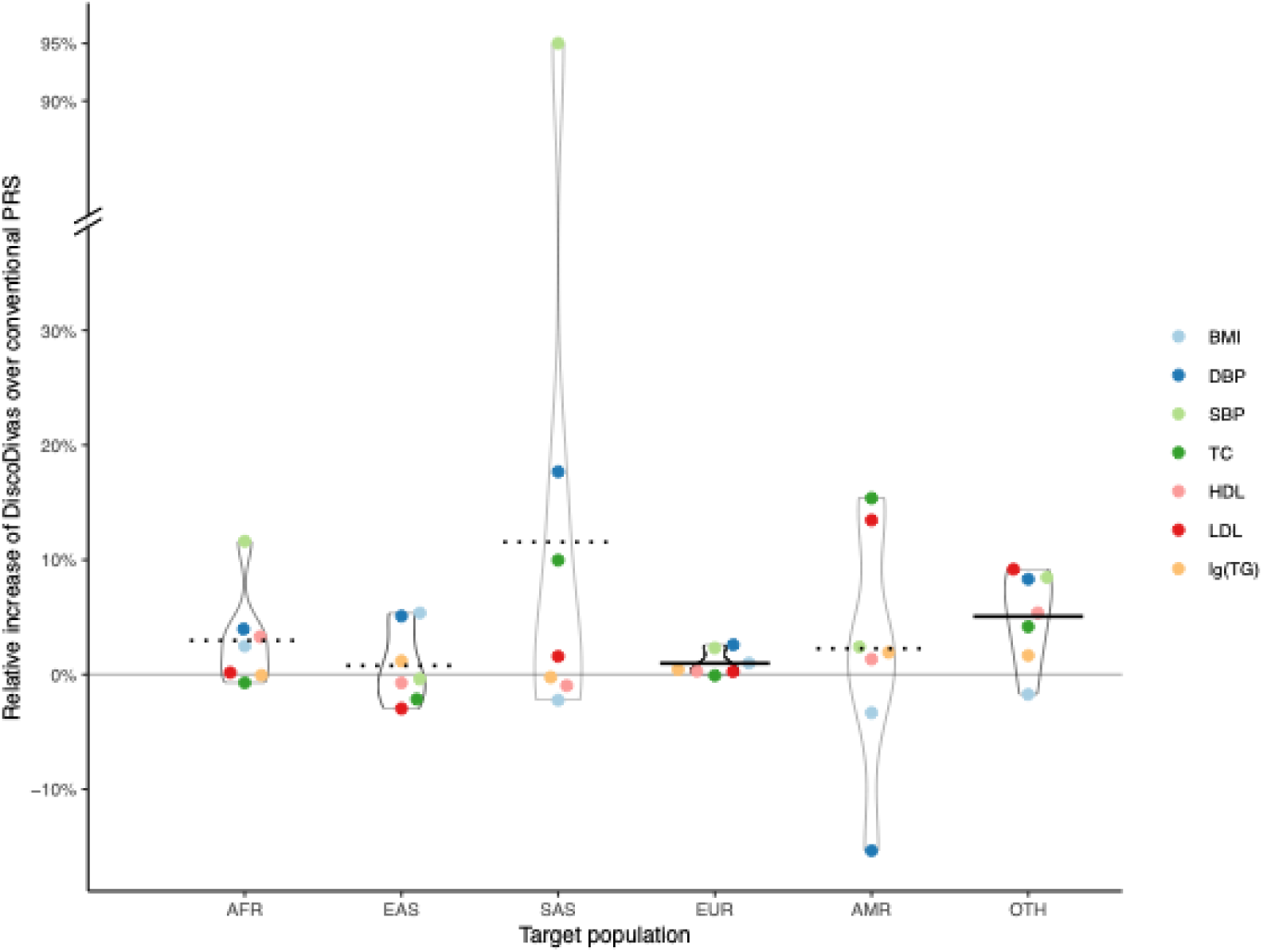
Relative R^2^ increase of DiscoDivas over the conventional PRS fine-tuned in a matched sample. The x-axis shows the population in which the PRS was tested. We used OTH as the fine-tuning dataset for the test in both OTH and AMR due to the absence of matched AMR training data. The horizontal bar shows the mean of relative increase, and the line-type of the bar indicates the *p-*value of paired t-test of DiscoDivas PRS R^2^ and conventional PRS R^2^, with the solid bar being *p-*value <0.05 and dotted bar being *p-*value>0.05.

For the binary traits coronary artery disease (CAD) and type 2 diabetes (DM2) (Figure 4), we used the AFR, EAS, EUR, SAS, AMR, and OTHsamples from All of US Research (AoU) as the fine-tuning data and tested in AFR, EAS, EUR, SAS, and OTH individuals in UKBB and AFR, EAS, EUR, SAS, and AMR individuals in MGBB. The DiscoDivas PRS were based on the PRS fine-tuned in AFR, EAS, EUR, and SAS and used the default assumption that the PRS fine-tuned from all the samples were of similar quality even though the sample sizes of both discovery GWAS and the fine-tuning samples were not balanced across different ancestries. AMR in UKBB was excluded because of the small sample size (N=669).

**Figure 4.**
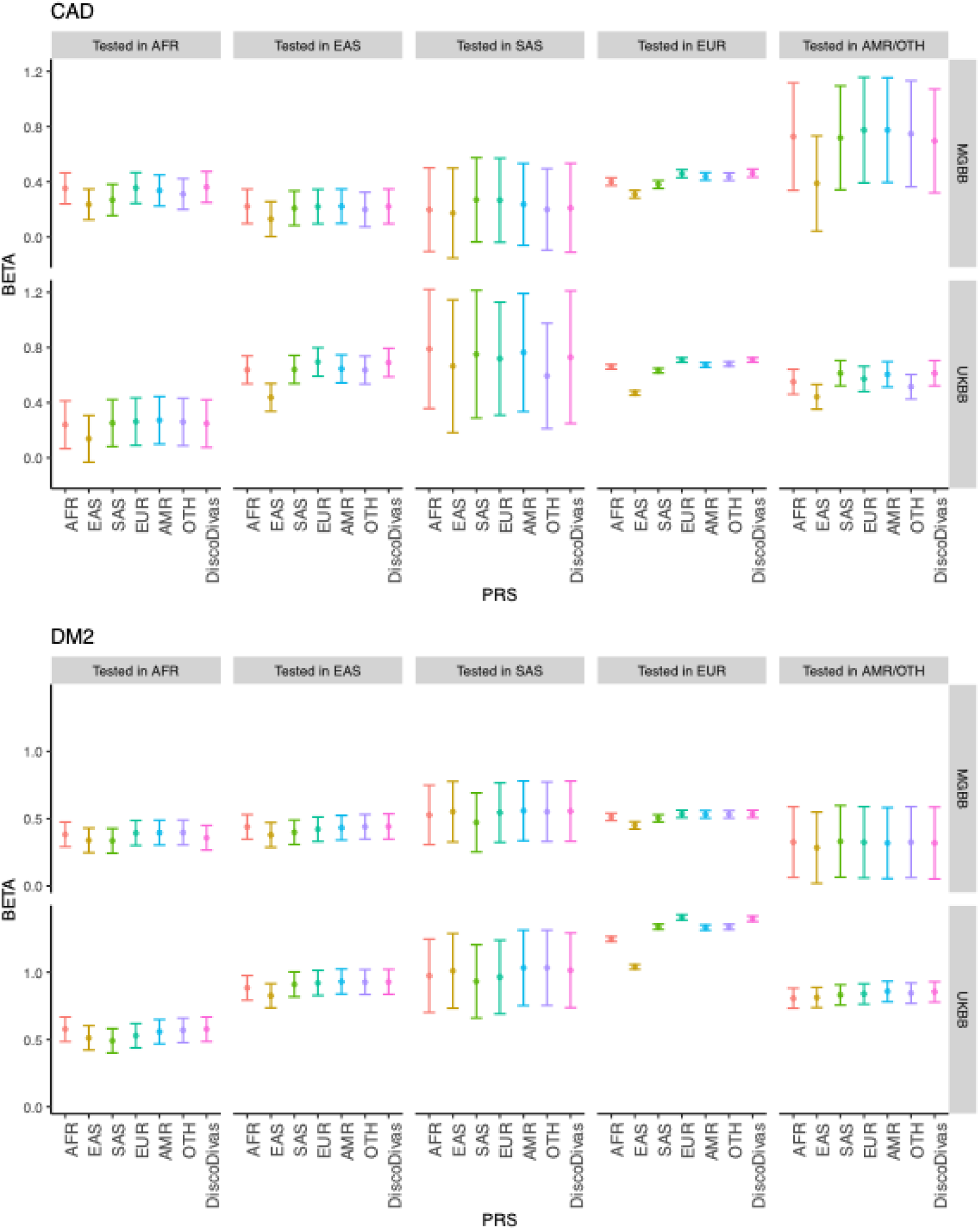
PRS performance for coronary artery disease (CAD) and type 2 diabetes (DM2) tested in UKBB and MGBB. The plot shows BETA, which is defined as ln(OR per SD) with the error bar showing 95% CI. The sub-panels show that population of the testing sample and the different colors show the method for generating the PRS, either fine-tuning in a single sample or combining the PRS using DiscoDivas.

The PRS fine-tuned in different single samples and the DiscoDivas PRS had similar performances. It also appeared that some of the fine-tuning sample could be underpowered: generally, we expect the PRS fine-tuned in the matched sample to perform the best in the testing samples, but PRS fine-tuned in larger fine-tuning data performed better than PRS fine-tuned in smaller fine-tuning data in general. For example, the PRS fine-tuned in EAS AoU data performed worse than other PRS in both MGBB and UKBB EAS data and had low accuracy in other testing data as well; the CAD PRS fine-tuned in EUR performed better than all the other PRS in all the testing data and the effective sample size of EUR CAD fine-tuning data was much larger than all the other fine-tuning data.

## Discussion

We propose a new method, DiscoDivas, to interpolate the PRS for diverse, especially admixed, ancestries with a generalized framework that does not required binning into discrete ancestries. Our results shows that the accuracy of DiscoDivas was comparable to or greater than the conventional method, i.e. fine-tuning using the matched population sample when available. In addition, when generating PRS for a wide range of ancestries, DiscoDivas did not require shifting from several sets of PRS models fine-tuned in discrete samples while remaining matched with the ancestry information. Additionally, DiscoDivas is a flexible framework that allows input PRS models to be generated using other advanced methods, further enhancing the final PRS performance across diverse genetic ancestries. Our method provides a new solution to generate PRS for underrepresented, generally admixed, populations and as well as generate a harmonized PRS model across different ancestries.

The performance of our method depends on the quality of both the discovery GWAS data and the fine-tuning data. As shown in the simulation test, discovery GWAS datasets that represent diverse ancestries with sufficient sample size will increase the accuracy of interpolated PRS generated by DiscoDivas. On the contrary, Eurocentric and underpowered discovery GWAS datasets would limit the advantage of DiscoDivas over the conventional PRS method. This might partly explain the limited advantage of DiscoDivas when predicting binary traits: the discovery GWAS datasets were highly Eurocentric and the GWAS, especially the non-European cohorts, could be more underpowered than quantitative trait GWAS. Furthermore, the PRS fine-tuned in fine-tuning datasets of insufficient sample size will be overfitted and cannot be used to fairly evaluate the performance of either the conventional PRS method or DiscoDivas. We aimed to address this issue by only using traits that 1) had effective sample sizes larger than 200 in all the fine-tuning samples, and 2) had high-quality phenotyping data in both the fine-tuning datasets and the testing datasets, However, Asian populations were largely under-represented in the current public biobanks: the effective sample size of many binary traits in EAS or SAS can be as small as <200 even in AoU, the most diverse and large-scale largely publicly-available biobank we had access to. This limited our choice for binary traits to only CAD and DM2.

The difficulty of gathering data for our research underscores the notion that non-European populations, both admixed and singe-ancestry populations, remain largely under-represented in the existing genetic data. Furthermore, some potential extensions of our method will not become possible until we collect more diverse and larger datasets. First, our method has not been designed nor tested for extrapolating data, e.g. generating PRS for continental African samples based on African American, European, and Asian samples. Even though it is mathematically plausible to alter our method to extrapolate the PRS, we lack data such as continental African samples to test the method. Secondly, we currently only consider the assumption that the most informative genome-wide PRS model shifts linearly in the PCA space. Although more complicated PRS interpolation, e.g. interpolation guided by local ancestry information ^24,33,37^, pathway-specific^39,40^ and annotation-guided^41^ PRS models and polynomial interpolation^42,43^, can possibly further improve the PRS accuracy, training such complicated models would require collecting much larger and more diverse datasets than the existing data. Finally, additional biological insights could be revealed by interpolating PRS if genetic data of all the involved diverse ancestries are of sufficient power. In this case, the differences between interpolated PRS and the PRS trained using the matched ancestry would indicate the population- or sample-specific factors absent in the interpolation model, e.g. population-specific genetic variance^44^, complicated population stratification involving cofounding factors^45,46^, sample/ancestry-specific modifiers like local adaptation^45^, gene x environment interactions^47^ or other factors that contribute to the genetic variant frequency or effect size in these samples/ ancestries.

DiscoDivas does not consider the local ancestry information, which improve PRS predictions in various research^24,33,37^, especially PRS prediction of newly admixed populations^38^. This may result in suboptimal performance in newly admixed populations. However, this design choice makes our method highly applicable and robust in settings where the input PRS model are fine-tuned using cohorts whose genetic ancestries deviate from those in local ancestry inference reference panels and where the local ancestry of the testing cohort is difficult to infer — for instance, due to deep or complex admixture events that have resulted in highly fragmented ancestry tracts^25,26^.

In conclusion, our method provides a new option to treat the ancestry information as a continuous variable and interpolate a harmonized PRS for diverse ancestries.

Notably, although our method was developed primarily to calculate PRS when the matched fine-tuning datasets were unavailable, our research showed that successfully interpolating PRS required sufficient input data and highlighted the need to collect genetic data for underrepresented populations. We believe that more diverse and larger data collected in the coming future will enable the development of new methods of interpolating PRS and the elucidation of the genetic basis of complex traits.

## Methods

### Methodological Details of DiscoDivas

DiscoDivas interpolates PRS of testing individuals of diverse ancestry according to the testing individual’s PRS calculated using the PRS model fine-tuned in a few single-ancestry fine-tuning datasets and genetic distance information. The pipeline consists of two parts: harmonizing the input PRS data and interpolating the PRS.

#### Data harmonization

To reduce the bias in the interpolation, the PRS and PCA information should be in a unified and harmonized scale. First, all the individuals in the fine-tuning datasets and the testing dataset are projected in the same PCA space based on balanced reference data covering the global genetic ancestry continuum. The reference data is essential to avoid skewed correlation between the genotypic similarity and the genetic distance, and to ensure that the PCA Euclidean distance can present consistent genetic distance. The results presented in this study were based on PCA calculated using pruned SNPs of 1000 Genomes^31^ samples. A detailed description of calculating PCA and Euclidean distance in this research is provided in the supplementary section entitled ‘Calculation of 1000 Genomes-based PCA and Euclidean distance.’

Second, all the PRS input should be transferred to a comparable scale. We regressed out the top 10 PCs from the PRS and then standardized the PRS residuals to mean = 0 and standard deviation = 1.

#### PRS interpolation

The overall mathematical model of DiscoDivas is a linear combination of PRS based on the PRS model fine-tuned in different fine-tuning datasets:

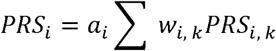

where *PRS_i, k_* is the normalized PRS of individual *i* trained in fine-tuning dataset *k; w_i, k_* is the interpolation coefficient which is a function of genetic distance between the individual *i* and dataset *k* and other factors. *a_i_* is a constant for individual *i*so that Σ*a_i_ w_i, k_* = 1.

The essential factor contributing to *w_i, k_* is the reciprocal of *D_i-k_*, the genetic distance between individual *I* and dataset *k*, so that the interpolation is basically linear. In the ideal situation where all the fine-tuning samples are independent and generate PRS of the same level of accuracy, the genetic distance between the testing individuals and fine-tuning datasets is the only contribution factor and we define:

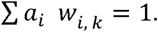

However, considering the more realistic scenarios where the fine-tuning samples may be correlated and the PRS trained from different fine-tuning datasets may be of different accuracies, we introduce two parameters:

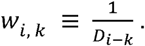

1. *d_k_*: tuning parameters based on the genetic distance / correlation between the training datasets.
2. *r_k_*: tuning parameter that represent the accuracy of the PRS fine-tuned in sample *k;*

so that the overall interpolation coefficient is

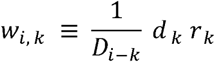

The final model of DiscoDivas is

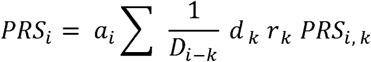

Here we propose the default method for calculating *d_k_* and *r_k_*: *d_k_* is based on the genetic distance matrix *G* in which each row and column samples *i* and *k*, with diagonal ones being zero :

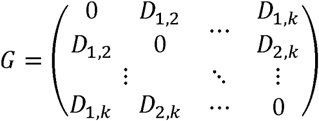

The shrinkage follows the similar principle of correcting marginal SNP effect size by inverse of the LD matrix, which captures the correlation structure among SNPs. Analogously, since genetic distance reflects the correlation between PRS models, its inverse can be used to inform the degree of adjustment needed when combining or interpolating PRS models across ancestries. The vector of shrinkage parameter *d_k_* is only derived from *G*^-1^:

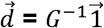

where 1⃗ is a vector of the same length as *d⃗* and with all the elements being 1. *r_k_* is based on the accuracy of the PRS fine-tuned in the sample k.

#### Different options to decide the value of *r_k_*

Under the additive assumption, the maximum accuracy of a PRS is achieved when the PRS R^2^ approximates narrow-sense heritability *h*^2^ and the PRS is saturated (namely, adding more samples in the Discovery GWAS would not further increase the PRS accuracy if other conditions remain the same). Previous research used percentage of heritability explained to present the accuracy of the PRS so that PRS predicting traits of different heritability and binary traits of different prevalence can be compared^48,49^. Here we recommend using the proportion of narrow-sense SNP heritability in the target sample explained by the PRS as the first choice of *r_k_*

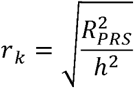

However, the accurate estimation of *h*^2^ requires sufficient sample size, which may accurately not always be available for the individual-level data. If *h*^2^ is unknown or cannot be estimated, we provided an alternative approach that approximates the PRS given that most of the PRS is far from saturation and the accuracy of the PRS is roughly linearly correlated with the sample size of the discovery GWAS *N_k_*, we combine the information from different samples in a similar way by combining Z scores in fixed-effect size meta-analysis:

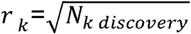

If the trait or genetic ancestry used in the discovery GWAS does not match that of the fine-tuning and testing datasets, which collectively referred to as the ’target data,’ the PRS accuracy also depends on the heritability of the trait in the discovery GWAS *h_k, discovery_*^2^ and the genetic correlation between the discovery GWAS trait and the target trait *rg_k discovery-target_*,

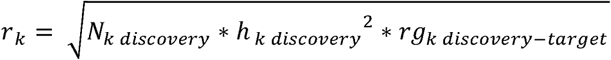

In a common and more ideal scenario where discovery data comes from multiple GWAS from different genetic ancestries and of decent statistical power for each genetic ancestry yet the heritability of the target trait in the fine-tuning sample is unknown or cannot be accurately estimated, the accuracy of the PRS fine-tuned in each single-ancestry fine-tuning sample is hard to estimate but is likely of similar accuracy. Therefore *r_k_* can be omitted or equivalently set to a default constant value of 1.

In the DiscoDivas script provided, the *r_k_*, is set to be the default value 1 unless defined by the user otherwise, *a_i_*, *D_i-k_*, *d_k_* is automatically calculated from the PCA information provided by the user.

### Data

#### UK Biobank

The UK Biobank (UKBB) is a volunteer sample of approximately 500,000 adults aged 40-69 upon enrollment living in the United Kingdom recruited since 2006^50^. UKBB data used in this research were first QC’ed with the following process: Remove the individuals meeting the criteria that indicate low genotype quality or contamination: 1) have missing genotype rate larger than 0.02; 2) have genotype-phenotype sex discordance; 3) are identified as having excess heterozygosity and missing rates; 4) are identified as putatively carrying sex chromosome configurations that are not either XX or XY; 5) appeared to have unreasonably large numbers of relatives. From the remaining samples, only one individual from a group of multiple individuals that are closer than 3^rd^-degree relatives was retained. 415,402 individuals were left after the QC. 390,037 were self-identified as EUR, 7,039 AFR, 8,652 non-Chinese Asian (ASN), 1430 Chinese (CHN) and 6572 unknown or not answered, and 1672 as admixed (MIX). The genetic ancestry referred from PC was largely correlated with the self-reported race, with 385,038 EUR, 7,450 AFR, 8,298 SAS, 2,163 EAS, 669 AMR and 11,784 other (OTH) or admixed.

In the PRS test, UKBB samples were grouped by their genetic ancestry (see section ‘Genetic ancestry inference’). The fine-tuning datasets for the single-ancestry populations (AFR, EAS, EUR and SAS) were based on 1300 randomly selected samples whose self-report ancestry matched with their genetic ancestry and the probability of random forest = 1. The fine-tuning dataset for admixed ancestry (OTH) is 1300 randomly selected samples of individuals of OTH genetic ancestry (see Supplementary Figure 3). AMR didn’t have its corresponding fine-tuning dataset due to its small sample size, and we used OTH fine-tuning datasets as a proxy since the two genetic ancestries had similar PCA. The remaining individuals of UKBB were used as testing data.

The quantitative trait of the UKBB samples was the first measurement collected after enrollment. The lipid trait measurement was adjusted for cholesterol-lowering medication by dividing TC by 0.8 and LDL by 0.7 as before^51^. Cases of coronary artery diseases (CAD) are defined using the definition described previously^24^; Cases of type 2 diabetes (T2D) were defined as meeting any of the followings: 1) ICD-10 of E11 or self-reported diagnosis of diabetes with available T2D first diagnosis date, or 2) HbA1C ≥ 6.5%, or 3) insulin use at enrollment, or 4) use of oral glucose-lowering medication excl. isolated Metformin prescription at enrollment; controls were defined as 1) without any diabetes or prediabetes, and 2) HbA1c < 5.7%. Cases of Type 1 Diabetes (1) ICD of E10 or self-report diagnosis of DM with first diagnosis date of type 1 diabetes; and 2) insulin initiation within 1 year of diagnosis and 3) insulin use at enrollment), prediabetes (without diabetes and HbA1c ≥ 5.7% and HbA1c < 6.5%) were excluded.

UKBB participants provided consent in accordance with the primary IRB protocol, and the Massachusetts General Hospital IRB approved the present secondary data analysis of the UKBB data under UKBB application 7089.

#### Mass General Brigham Biobank

The Mass General Brigham Biobank (MGBB) is a volunteer sample of approximately 142,000 participants receiving medical care in the Mass General Brigham health care system recruited starting 2010. 53,306 MGBB participants underwent genotyping via Illumina Global Screening Array (Illumina, CA). MGBB genotype data was quality controlled, imputed and assigned one of the populations AFR, AMR, EAS, EUR, SAS using K-nearest neighbor model as described previously^52^. The phenotype data of CAD and diabetes are drawn from PheCodes based on International Classification of Diseases codes, Ninth (ICD9)110 and Tenth (ICD10) revisions, from the EHR as described previously ^52^. MGBB participants provided consent in accordance with the primary IRB protocol, and the Massachusetts General Hospital IRB approved the present secondary data analysis.

#### All of Us Research Program

The *All of Us* (AoU) Research Program is a volunteer sample of more than one million United States residents recruited starting 2016. AoU aims to engage communities previously underrepresented in biomedical research in the United States and beyond^53^. In the present analysis, genetic data from the v7 245,394 participants who were genotyped using short read whole genome sequencing (srWGS) data. Hapmap3 SNPs were extracted for the callset with the threshold of (AF) > 1% or population-specific allele count (AC) > 100. Related individuals were pruned according to the information provided by AoU. Due to the inclusive data collection, we didn’t exclude individuals whose self-report gender were different with their assigned sex at birth and used the combination of self-report gender and assigned sex as one of the covariates. The predicted ancestry information was provided by AoU^54^. The phenotypes were defined as described in previous research by Buu *et al*^32^.

#### Simulated data

The simulated GWAS summary statistics were based on simulated genotype data based on 1000 Genomes reference^6^. Only Hapmap3 SNPs were included in the simulation. Causal SNPs were randomly selected from the Hapmap3 SNPs and simulated per allele effect size following normal distribution. The non-genetic factor was set to follow the same normal distribution across populations. The ladder of causal SNP number was 100, 1000, 3000, 10000 and the heritability in each of the population was 0.6. The causal SNP effect size was simulated as either constant across populations or varying linearly in the PCA space.

The phenotype is the sum of genetic burden and non-genetic factor:

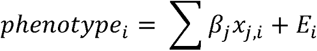

where the *phenotype_i_* and *E_i_* were the phenotype and non-genetic factor of individual *i;β_j_* was the effect size of causal SNP *j*, and *x_j,i_* was the number of risk alleles of individual *i* in SNP *j*.

We used the PLINK2^28^ to calculate the genetic burden based on the simulated causal SNPs and effect size and used R to simulate the non-genetic factors by generating a normal distribution, scale the genetic burden and non-genetic factor, and generate a phenotype of heritability set to be 0.6. We used up to 100k individuals per population to generate the summary statistical GWAS as the discovery data for the PRS test.

The rest simulated data were left out for the fine-tuning and testing datasets. The summary statistics GWAS were generated based on the simulated genotype data and phenotype data using the ‘--glm’ function of PLINK2.

In addition to the completely simulated data, we generated more realistic fine-tuning and testing datasets of a wider genetic ancestry range by using the QC’ed genotype data from UKBB described in the section ‘Biobank data.’ We simulated the genetic burden, non-genetic factor, and phenotype based on the real-life UKBB genotype data with the same pipeline and parameters. A more detailed description of simulating the data were given in section ‘Generating data for simulation analysis’ in the supplementary text.

## Supporting information

Supplementary Figures with legend

## Data Availability

The access to biobank data (UK Biobank, Mass General Brigham Biobank, and All of US Research Program) were gained upon application. The simulated genotype data based on 1000 Genomes were downloaded from https://dataverse.harvard.edu/dataset.xhtml?persistentId=doi:10.7910/DVN/COXHAP; The resource of summary statistics GWAS data used to generate PRS were given in the supplementary file. The 1000 Genome raw reference genotype data were downloaded from https://cncr.nl/research/magma/.

## Code Availability

The scripts of running DiscoDivas and other supporting files can be found at https://github.com/YunfengRuan/DiscoDivas; PRS-CS: https://github.com/getian107/PRScs; LDpred2: https://privefl.github.io/bigsnpr/articles/LDpred2; PLINK2: https://www.cog-genomics.org/plink/2.0/.

## Acknowledgements

We thank the participants and staff of the UK Biobank (application 7089), Mass General Brigham Biobank, and *All of Us* Research Program. We thank Ying Wang, Paul O’Reilly, Raymond Walters for helpful discussion and advice. P.N. is supported by NHGRI U01HG011719 and NHLBI R01HL127564. N.C is supported by NHGRI U01HG011719 and R01HG010480. A.P.P. is supported by NHLBI K08HL168238. S.M.C. is supported by NHLBI K099HL177340.

## Author Contributions

Y.R. and P.N. designed the project; Y.R. developed the statistical methods and programmed the code for DiscoDivas. A.P curated the summary statistical GWAS. R.B, S.K, L.H, B.T, S.M.C., and W.H participated in application for the access to the Biobank data and the data curation. Y.R and R.B performed the data analysis. R.B, B.T, H.Z, and N.C contributed to the method development. Y.R. and P.N wrote the manuscript. A.P. and N.C. provided critical revision for the manuscript. All the authors reviewed and approved the final version of the manuscript.

