## supplementary text for "DiscoDivas: Leveraging genetic ancestry continuum information to interpolate PRS for admixed populations"

### Supplementary Methods

#### Calculation of 1000 Genomes-based Principal components of ancestry and Euclidean distance

We use 1000 Genomes as the reference panel for PCA calculation. The PCA should be based on SNPs that are constantly included in as many samples as possible to enable the use of wide-ranging discovery GWAS and fine-tuning datasets. We started with the Hapmap3 SNPs for this set of SNPs, which has been widely used as a subset of SNPs that approximates the feature of genome-wide common SNPs in many recent studies that involve multi-ancestry prediction ^1–4^. We further filtered for the SNPs likely to be frequently genotyped or imputed with relatively high quality by most samples based on the 1000 Genome data: Hapmap3 SNPs were first extracted from the five super-populations, Africans (AFR), Admixed Americans (AMR), East Asians (EAS), Europeans (EUR) and South Asians (SAS) of the 1000 Genomes. Secondly, SNPs described as the following were excluded: 1) of minor allele frequency lower than 1% in any of the super-population, 2) of minor allele frequency lower than 5% in the combined 1000 Genomes data, and 3) in the long-range LD region (25Mb – 35Mb by hg19 assembly on chromosome 6 and 7Mb – 13Mb on chromosome 8). To calculating the PCA loading, the QC’ed SNPs of the five super-populations were merged then pruned using the PLINK2 function “indep-pairwise” with the parameter “200 100 0.1” - namely the pruning was performed using window size = 200kb, step size = 100, and phased-hardcall-r^2^= 0.1. The principal components and the SNP loadings are calculated using PLINK2 function “pca” with the parameter “allele-wts” based on the pruned SNPs.

Based on the protocol suggested on the PLINK2 website (<https://www.cog-genomics.org/plink/2.0/score#pca_project>), we projected samples for fine-tuning and PRS testing into the PCA space as describe above by calculation the linear score, i.e. the sum of alternative alleles weighted by the SNP effect size, using the PLINK2 function “score” with the SNP loadings as effect size. The original online protocol suggested linear score should be first scaled to standard variation and then rescaled by multiplying the square root of eigenvalue. However, the actual standard deviation of a sample in the same PCA space varies with the homogeneity and the ancestry of the sample. Forcing the PCA of all the samples to have the same standard deviation will cause inconsistent scaling when the samples can be of different ancestries. Therefore, we directly calculated the PCA from sum basic linear score based on the SNP loadings as generated above without any further scaling. The PCA in this study was the sum basic linear score calculated using the PLINK2 function “score” with the parameter “cols=+scoresums'”. For large samples whose genotype data were divided into per-chromosome files, the same commands were used to calculate per-chromosome score and the genome-wide score was the sum of the score of all the autosomes.

In DiscoDivas’ default setting, the genetic distance between two individuals is defined as the Euclidean distance between the PCA of the two individuals. When the genetic distance calculation involves a sample of multiple individuals, we use the median point to present the whole sample.

We also explored the relationship between number of PCs included in the calculation and the Euclidean distance calculated (Supplementary Figure 9) and the distance calculated converged when the number of PCs was larger than 6 in our tests. In our analysis we use the top 10 PCs to calculate the PCs.

#### Genetic ancestry inference

We noticed that the protocol of generating top PCs for ancestry inferences varied in previous publications. In our pilot test (see Supplementary Results subsection entitled ‘Pilot test of generating PCA based on less QC’ed SNPs’), we compared the ancestry inference based on Hapmap3 SNP without any QC and found the result to be highly correlated. We used the same set of PCs based on QC’ed SNP as described in section ‘Calculation of 1000 Genomes-based PCA and Euclidean distance’ for both genetic ancestry inference and Euclidean distance calculation for data consistency.

Random forest model of 100 trees was trained based on the 1000 Genome data. The out-of-bag estimate of error rate stabilize at the level of 0.28% after the number of PCs passed 5. We used the model using the top 6 PCs to infer the genetic ancestry of UK Biobank individuals and the Mass General Brigham Biobank individuals. The genetic ancestry of an individual was assigned to any of the five ancestries represented in the 1000 Genomes reference data, i.e. AFR, AMR, EAS, EUR and SAS, if the highest probability of an individual belonging to that ancestry passed a threshold. If none of the ancestries had a probability above the threshold, the individuals were assigned as other (OTH), which indicated that the individual was of admixed ancestries. With the consideration of the sample size and confirmed by visual inspection, the threshold of probability for UK Biobank and the Mass General Brigham Biobank was 0.9 and 0.8 respectively.

#### Methodological Details of PRS Construction Using a Single Fine-tuning Dataset

The PRS generated with single-tuning sample were derived from multiple GWAS conducted in different populations. For simulation test, the GWAS were generated based on simulated genotype and phenotype as described in Supplementary Methods subsection entitled ‘Generating data for simulation analysis’; for empirical test, the GWAS were listed in Supplementary Table 1.

GWAS data were first processed with PRS-CS or LDpred2 to generate adjusted SNP effect size. In our pilot tests, we observed that LDpred2 had difficulty converging for certain simulated traits. Additionally, for real-world traits, the best-performing PRS models generated by PRS-CS and LDpred2 showed comparable performance, with LDpred2 typically performing slightly better. To reduce computational cost, we therefore chose to use PRS-CS exclusively for simulated data and LDpred2 exclusively for real-world data. The procedures for running both methods are described as follows:" Hapmap3 SNPs were first extracted from each GWAS as the input for the PRS method. The PRS methods were performed using default parameters: For PRS-CS, parameters of the prior distribution were set to phi = 1, 10^-2^, 10^-4^, 10^-6^, a = 1, b = 0.5 and the parameters of Markov Chain Monte Carlo (MCMC) were total number of MCMC iterations = 1000, number of burn-in = 500, thinning factor of the Markov chain = 5. For LDpred2, the parameters were the default set as in previous research^5^: proportion of variants assumed to be causal was 1.0 × 10^−4^, 1.8 × 10^−4^, 3.2 × 10^−4^, 5.6 × 10^−4^, 1.0 × 10^−3^, 1.8 × 10^−3^, 3.2 × 10^−3^, 5.6 × 10^−3^, 1.0 × 10^−2^, 1.8 × 10^−2^, 3.2 × 10^−2^, 5.6 × 10^−2^, 1.0 × 10^−1^, 1.8 × 10^−1^, 3.2 × 10^−1^, 5.6 × 10^−1^ and 1, the scale of heritability was  0.7, 1 and 1.4 times of the estimated heritability, with options of whether allowing a sparse output or not.

Each set of parameters generated a corresponding set of adjusted SNP effect size, which were then used to calculate PRS in the fine-tuning samples. The most predictive PRS for each GWAS was selected based on a linear or logistic regression model predicting the phenotype using the PRS and adjusting for top 20 PCs, age, sex and genotyping batch for biobank empirical analyses, and adjusting for only top 20 PCs for simulated analyses. When analyzing All of Us data, the combination of assigned sex and self-reported gender, rather than assigned sex alone, was used as a covariant. This approach maximizes the sample size and leverages the inclusivity of the data collection.

To generate the final PRS model, multiple top-performing PRS based on each GWAS were combined through a linear or logistic regression in the fine-tuning sample. Final adjusted PRS models were a linear combination of the top SNP models from each GWAS, weighted by the regression coefficients. These combined PRS models were subsequently used to calculate PRS in the testing sample.

Conventionally, when fine-tuning PRS for a testing sample, it is ideal to use a sample from a matched or similar population. If such a sample is unavailable, the PRS is fine-tuned using any available sample, which is often from individuals of European ancestry.

Supplementary Table 1: Summary of discovery GWAS for empirical PRS testing. BMI: body-mass index; HDL: high-density lipoprotein cholesterol; LDL: low-density lipoprotein cholesterol; TC: total cholesterol; TG: triglycerides; SBP: systolic blood pressure; DBP: diastolic blood pressure; CAD: coronary artery disease (CAD); DM2: type 2 diabetes mellitus.

| Trait | Name | Dominant Ancestry | N Cases | N Controls | ref |
| --- | --- | --- | --- | --- | --- |
| CAD | CARDIO- GRAM plusC4D no UKBB | EUR | 86,847 | 417,789 | ^6^ |
|  | BBJ | EAS | 29,319 | 183,134 | ^7^ |
|  | Genes&Health | SAS | 996 | 16,352 | ^8^ |
|  | FinnGen | EUR | 33,628 | 275,526 | ^9^ |
|  | MVP | EUR | 95,151 | 197,287 | ^10^ |
|  | MVP | AFR | 17,202 | 59,507 | ^10^ |
|  | MVP | HISP | 6,378 | 24,270 | ^10^ |
| DM2 | Diamante/MVP | EUR | 148,726 | 965,732 | ^11^ |
|  | MVP | AFR | 24,646 | 31,446 | ^11^ |
|  | MVP | HISP | 8,616 | 11,829 | ^11^ |
|  | Diamante | AFR | 15,487 | 23,709 | ^12^ |
|  | Diamante | SAS | 16,540 | 32,952 | ^12^ |
|  | AGEN T2D | EAS | 77,418 | 356,122 | ^13^ |
|  | FinnGen | EUR | 49,303 | 255,466 | ^9^ |
|  | Genes&Health | SAS | 9,044 | 12,066 | ^14^ |
| BMI | GIANT | EUR | 339,224 | | ^15^ |
|  | Genes&Health | SAS | 13,926 | | ^8^ |
|  | ALSPAC | EUR | 5,766 | | ^16^ |
|  | PAGE | HISP | 49,839 | | ^17^ |
|  | BBJ | EAS | 163,835 | | ^7^ |
| DBP | MVP | EUR | 249,262 | | ^18^ |
|  | Genes&Health | SAS | 15,908 | | ^8^ |
|  | PAGE | HISP | 35,433 | | ^17^ |
|  | BBJ | EAS | 145,515 | | ^7^ |
| SBP | MVP | EUR | 249,262 | | ^18^ |
|  | Genes&Health | SAS | 15,908 | | ^8^ |
|  | PAGE | HISP | 35,433 | | ^17^ |
|  | ALSPAC | EUR | 5,751 | | ^16^ |
|  | BBJ | EAS | 145,505 | | ^7^ |
| LDL-C | GLGC (without UKBB) | EUR | 842,660 | | ^19^ |
|  | GLGC (without UKBB) | AFR | 87,760 | | ^19^ |
|  | GLGC (without UKBB) | SAS | 33,658 | | ^19^ |
|  | MVP | EUR | 215,551 | | ^20^ |
|  | MVP | AFR | 57,332 | | ^20^ |
|  | MVP | HISP | 24,742 | | ^20^ |
|  | BBJ | EAS | 72,866 | | ^7^ |
|  | Genes&Health | SAS | 10,939 | | ^8^ |
|  | ALSPAC | EUR | 5,766 | | ^16^ |
| HDL-C | GLGC (without UKBB) | EUR | 888,226 | | ^19^ |
|  | GLGC (without UKBB) | AFR | 90,805 | | ^19^ |
|  | GLGC (without UKBB) | SAS | 33,953 | | ^19^ |
|  | MVP | EUR | 215,551 | | ^20^ |
|  | MVP | AFR | 57,332 | | ^20^ |
|  | MVP | HISP | 23,946 | | ^20^ |
|  | BBJ | EAS | 74,970 | | ^7^ |
|  | Genes&Health | SAS | 11,316 | | ^8^ |
|  | ALSPAC | EUR | 5,766 | | ^16^ |
| TC | GLGC (without UKBB) | EUR | 930,671 | | ^19^ |
|  | GLGC (without UKBB) | AFR | 92,555 | | ^19^ |
|  | GLGC (without UKBB) | SAS | 34,135 | | ^19^ |
|  | MVP | EUR | 215,551 | | ^20^ |
|  | MVP | AFR | 57,332 | | ^20^ |
|  | MVP | HISP | 24,743 | | ^20^ |
|  | BBJ | EAS | 135,808 | | ^7^ |
|  | Genes&Health | SAS | 13,113 | | ^8^ |
| TG | GLGC (without UKBB) | EUR | 864,239 | | ^19^ |
|  | GLGC (without UKBB) | AFR | 89,468 | | ^19^ |
|  | GLGC (without UKBB) | SAS | 34,023 | | ^19^ |
|  | MVP | EUR | 215,551 | | ^20^ |
|  | MVP | AFR | 57332 | | ^20^ |
|  | MVP | HISP | 24,063 | | ^20^ |
|  | BBJ | EAS | 111,667 | | ^7^ |
|  | Genes&Health | SAS | 11,125 | | ^8^ |
|  | ALSPAC | EUR | 5,766 | | ^16^ |

Supplementary Table 2: The binary trait effective sample sizes and total sample sizes of different genetic ancestry groups in All of Us, UK Biobank, and Mass General Brigham Biobank.

| Trait | Genetic Ancestry | AoU | | UKBB | | MGBB | |
| --- | --- | --- | --- | --- | --- | --- | --- |
|  |  | N_eff | N total | N_eff | N_total | N_eff | N_total |
| CAD | AFR | 9141 | 38916 | 572 | 7110 | 1461 | 2714 |
|  | EAS | 580 | 3813 | 133 | 2130 | 218 | 1008 |
|  | EUR | 37026 | 98609 | 64099 | 356166 | 23446 | 41941 |
|  | SAS | 437 | 1809 | 2476 | 7921 | 195 | 569 |
|  | AMR | 5654 | 1799 | 47 | 664 | 1173 | 3511 |
|  | Other | 5193 | 15060 | 1826 | 11203 | - | - |
| DM2 | AFR | 21782 | 52338 | 2636 | 4162 | 2455 | 2714 |
|  | EAS | 1089 | 5333 | 408 | 1533 | 422 | 1008 |
|  | EUR | 38838 | 122113 | 61646 | 319079 | 24609 | 41941 |
|  | SAS | 701 | 2328 | 4440 | 5643 | 319 | 569 |
|  | AMR | 15582 | 39990 | 102 | 527 | 2543 | 3511 |
|  | OTH | 5509 | 19008 | 2663 | 9132 | - | - |

Supplementary Table 3: Summary description of datasets used in each test.

| Datasets | Simulation Test | Quantitative Trait Test | Binary Trait Test |
| --- | --- | --- | --- |
| Discovery | Summary statistics GWAS generated from simulated genotype data based on 1000 Genome reference and simulated phenotype of sample size of 100k or 40k | Published summary statistics GWAS | Published summary statistics GWAS |
| Fine-tuning | For the main test: simulated data generated from genotype of 1300 UKBB individuals per population that are close to the 1000 Genome reference and simulated phenotype.  For the sensitivity test:  10k individual-level data generated from simulated genotype data based on 1000 Genome reference and simulated phenotype | Empirical data of 1300 UKBB individuals per population that are close to the 1000 Genomes reference | *All of Us* |
| Testing | simulated individual-level data generated from the remaining UKBB genotype data and simulated phenotype | The remaining UKBB individuals | UKBB and MGBB |

#### Generating data for simulation analysis

#### Overview of the simulation process

In the simulation analysis, we assumed that the phenotype is the sum of genetic burden and non-genetic factor:

$${phenotype}_{i}= \sum\beta_{j}x_{j,i}+E_{i}$$

where the ${phenotype}_{i}$ and $E_{i}$ were the phenotype and non-genetic factor of individual $i$; $\beta_{j}$ was the effect size of causal SNP $j$, and $x_{j,i}$ was the number of risk alleles of individual $i$ in SNP $j$. The genetic burden $\sum\beta_{j}x_{j,i}$ was the sum of causal SNPs weighted by the effect size.

The causal SNPs were randomly selected, and the value of causal SNP effect size and non-genetic factors were random numbers drawn from normal distributions. The process of selecting causal SNPs, assigning effect size, simulating phenotype data, and the downstream GWAS and PRS analysis was repeated 20-fold.

##### Genotype data used for simulation

To generate the simulated GWAS summary statistics, the genotype data was generated by Zhang et al^2^ and downloaded from <https://dataverse.harvard.edu/dataverse/multiancestry>. Only Hapmap3 SNPs (1.4 million) were included in the simulation. In addition to the completely simulated data, we generated more realistic fine-tuning and testing datasets of a wider ancestry range by using the QC’ed genotype data from UKBB described in the section ‘Biobank data’ of the main text. While we used all the non-European testing data, the EUR testing dataset was down-sampled to 10,000 for the simulation test to reduce the computation burden. 1300 individual from each population were used as the fine-tuning data and the rest were used as testing data. The simulated data based on UKBB genotype data were used as fine-tuning and testing data in the main test and the left-out completely simulated data were used as fine-tuning data in the sensitivity test.

##### Simulation of individual-level phenotype

We simulated phenotype based on simulated genotype and the real-life UKBB genotype data with the same pipeline and parameters.

First, causal SNPs were randomly selected from the Hapmap3 with the ladder of causal SNP number being 100, 1000, 3000, 10000. To simulate traits with causal SNP effect sizes constant across different ancestry, the genetic burden of individuals of different ancestries were calculated using the same set of per-allele SNP effect size drown from normal distribution. To simulate phenotypes based on UKBB genotype data for the scenarios of causal SNP effect size varying linearly in the PCA space with UKBB genotype data, we first simulated the causal SNP effect size of genetic ancestry represented by the median point of the different superpopulations of 1000 Genomes individuals in the PCA space. The effect size of different ancestries followed multivariate normal distribution with the covariance matrix being:

|  | EUR | SAS and EAS | AFR |
| --- | --- | --- | --- |
| EUR | 1 | 0.7 | 0.4 |
| SAS and EAS | 0.7 | 1 | 0.7 |
| AFR | 0.4 | 0.7 | 1 |

Similar to the principle that PRS models can be interpolated is equivalent to PRS can be interpolated, causal SNP effect size varies linearly in the PCA space and, therefore, can be interpolated is equivalent to genetic burden can be interpolated. The genetic burden of an individual is the weighted sum of what the genetic burden could be based on the simulated SNP effect size of the median point of different populations of 1000 Genomes, with the combination coefficient proportional to the reciprocal of the PCA distance.
We assumed that the non-genetic factor of individuals across different ancestries could be summed up as a quantitative variable independently drawn from the same normal distribution.

Causal SNPs were randomly selected using R function ‘sample’ and the effect sizes were simulated using R function ‘rnorm’ for normal distribution and ‘mvrnorm’ for multivariate normal distribution. The genetic burden was calculated using the PLINK2^28^ based on the simulated causal SNPs and effect size. Simulation of non-genetic factors, scaling of the genetic burden and non-genetic factor to generate a phenotype of heritability set to be 0.6 were performed with R. From the data based on simulated genotype data, we used up to 100k individuals per population to generate the summary statistical GWAS as the discovery data for the PRS test. The remaining data were left out for the fine-tuning and testing datasets

### Supplementary Result

#### Pilot test of generating PCA based on less QC’ed SNPs

We attempted two ways to generate PCA for UKBB individuals and compare if the PCA would influence the Random Forest classification results.

In addition to the PCA described in the Methods section, we generated a set of alternative PCA based on less-QC’ed SNPs. We extracted Hapmap3 SNPs from the 1000 Genome samples without any QC and used the same parameters to prune the genotype data and calculate the PCA loadings. The same process of projecting UKBB individuals into the PCA space and classifying individuals based on the PCA using Random Forest was performed. The PCA plot based on the two different SNPs demonstrated a significant resemblance in their patterns and trends, and the classification results were highly robust (supplementary table 1).

Supplementary Table 4: Comparison of classification results based on different PCA.

|  |  | Original classification result | | | | | |
| --- | --- | --- | --- | --- | --- | --- | --- |
|  | Genetic ancestry | AFR | AMR | EAS | EUR | SAS | OTH |
| Alternative classification result | AFR | 7308 | 0 | 0 | 0 | 0 | 142 |
|  | AMR | 0 | 613 | 0 | 0 | 0 | 56 |
|  | EAS | 0 | 0 | 1854 | 0 | 0 | 309 |
|  | EUR | 0 | 0 | 0 | 382901 | 0 | 2137 |
|  | SAS | 0 | 0 | 0 | 0 | 8121 | 177 |
|  | OTH | 62 | 31 | 7 | 381 | 19 | 11284 |

#### Sensitivity tests on using minor missing information or alternative choices of fine-tuning data

Since the quality of fine-tuning data is essential to the performance of DiscoDivas, we evaluated the influence of minor missing information or alternative choices of fine-tuning data with the following tests:

First, we considered the possible scenario where PRS models for different ancestries are provided from a publication but key detailed information about the fine-tuning data was not fully available, especially the PCA information of the fine-tuning datasets. A convenient approximation of the PCA of fine-tuning datasets is the median PCA value of the 1000 Genome^21^ individuals of a certain ancestry or “superpopulation.” Based on the simulation test as mentioned above, we tested the influence of replacing the actual PCA of the UKBB fine-tuning datasets with the 1000 Genome approximate on the 1) PCA Euclidean distance, 2) combination coefficient for interpolation, and 3) the PCA accuracy.

Since the UKBB individuals were selected to be included in fine-tuning datasets based on the PCA information of the 1000 Genomes, the PCA distribution of UKBB fine-tuning datasets closely aligned with that of the 1000 Genomes reference (Supplementary Figure 3). PCA distances between the testing individuals and the median point of fine-tuning datasets based on the actual UKBB fine-tuning data were highly correlated with the PCA distances based on the 1000 Genomes approximation for all the testing ancestry groups AFR, AMR, EAS, EUR, SAS, and OTH, with the data residing within the highly overlapped intervals and the Pearson correlation of individual datapoints close to 1 (Supplementary Figure 4). The combination coefficients were calculated assuming that the PRS models fine-tuned in the four fine-tuning datasets, i.e. AFR, EAS, EUR, and SAS, were of equal quality. According to the formula in Method session, $w_{i, k}$, the interpolation coefficient of the PRS based on the PRS model fine-tuned in fine-tuning sample $k$ for individual $i$ , should be $w_{i, k} \equiv\frac{1}{D_{i-k}} d_{k}$, with the $D_{i-k}$ being the PCA distance between individual $i$ and the median point of sample $k$; $d_{k}$ being the adjustment coefficient for PRS fine-tuned in sample $k$ based on the distance between the fine-tuning samples. $w_{i, k}$ was compared between the two scenarios of using the actual UKBB fine-tuning datasets versus using the 1000 Genomes approximate. The correlation of the combination coefficients was lower than the correlation of the PCA distance, especially for SAS testing individuals. However, each combination coefficient remained in almost the same range and the PRS fine-tuned in the SAS sample still had the highest interpolation coefficients (Supplementary Figure 5). When testing with the simulated data, the PRS R^2^ had almost identical distribution and Pearson correlation > 0.99 with the R^2^ of PRS based on the actual PCA information in all the simulated scenarios including when the causal SNP effect size varied with the ancestries (Supplementary Figure 6). The high similarity of the PRS accuracy despite the difference in combination coefficients might partly result from the correlation of the PRS fine-tuned in different samples and the constant interpolation coefficient range.

In addition, we tested if the results of DiscoDivas would remain robust for admixed individuals when using a different set of fine-tuning datasets. In addition to the primary simulation test where the fine-tuning datasets were simulated data based on UKBB genotypes, PRS were fine-tuned with the 10,000 left-out simulated datasets that were independent from the discovery GWAS while other parts of the analysis pipeline remained constant. The simulated AMR dataset was used to fine-tune PRS for AMR and OTH testing samples for the conventional method. The PRS R^2^ based on the two sets of fine-tuning datasets were compared in the scenarios where the discovery GWAS was based on 100k AFR, EAS, EUR, and SAS, and where the causal SNP effect sizes were constant across different ancestries. The correlation of PRS R^2^ based on UKBB-based fine-tuning datasets and purely simulated fine-tuning datasets of DiscoDivas was larger than 0.99 in at scenarios, much higher than that of conventional PRS method, which ranged from 0.73 to 0.98 (Supplementary Figure 7). The advantage of DiscoDivas over the conventional PRS method showed a similar pattern (Supplementary Figure 8)

#### Test the performance of DiscoDivas based on single-population GWAS using simulated data.

In this study, we mainly used the PRS based on all the available GWAS in each scenario as the input of DiscoDivas aligning with current practices to maximize the final PRS accuracy when generating PRS in empirical situations. To illustrate the possible performance of DiscoDivas under less ideal situations where input PRS was based on different sets of discovery GWAS and fine-tuned separately in each dataset, we altered the simulation test in the main text to model this type of situation. We only used one matched single-population discovery GWAS to generate each input PRS of DiscoDivas (Supplementary figure 10). The parameter representing the input PRS quality in this scenario was defined as $r_{k}$=$\sqrt{N_{k discovery}}$. We compared the performances of (1) the single-population PRS that performed the best in the corresponding fine-tuning data, (2) DiscoDivas PRS based on single-population PRS, (3) multi-population PRS fine-tuned in the matched population, and (4) DiscoDivas PRS based on multi-population PRS, with the simulated data based on admixed UKBB genotype. All the three methods based on GWAS from multiple populations had much better performance than PRS based on single-population GWAS as expected. Notably the three multi-population PRS was the linear combination of the PRS based on the same set of GWAS but the difference in optimizing the PRS could lead to a numerically marginal yet statistically significant difference. Despite using PRS based on different sets of discovery GWAS as described in this test is less optimal than utilizing PRS derived from all available GWAS as described in the main text, it remains a viable approach to improving PRS accuracy for admixed populations. This method offers an advantage over using PRS fine-tuned in an unmatched single-population cohort and is particularly relevant when data access limitations prevent fine-tuning with all available GWAS. (Supplementary Figure 11).

#### Sensitivity test on using other PCA software

Previous research has shown that the PLINK2 might produce inaccurate principal component of ancestry^22^. Therefore, we compare the PC calculated using PLINK2 and the corresponding Euclidean distance with those calculated using bigsnpr, a method that performed better in the previous research.

We used the same subset of SNPs of the 1000 Genomes individuals as the main analysis as the input of bigsnpr and calculated the PCA loading using the default setting. We projected UKBB samples using the same PLINK2 function as used in the main analysis based on the PC loadings calculated using PLINK2 and the PC loadings calculated using bigsnpr and compare the absolute value of correlation between the 2 sets of PC since the sign of the correlation does not influence the downstream Euclidean distance comparison. The top 9 PCs are highly correlated with the Pearson correlation > 0.9 (supplementary figure 12). We then compare the Euclidean distance between the testing individuals of the six different genetic ancestries (AFR, EAS, EUR, SAS, AMR and OTH) to the median point of the four fine-tuning subsets (AFR, EAS, EUR and SAS) (supplementary table 5), resulting in 24 combination of testing individual and fine-tuning subset for comparison. In all the comparison, the correlation between the two sets of Euclidean distance were close to 1 and the ratio of the numeric value of Euclidean distance was constant across all the combination with a standard deviation of only ~3% of the mean value.

Supplementary Table 5: Pearson correlation and Euclidean distance ratios between testing individuals and the median point of UKBB fine-tuning cohorts, calculated using principal components from PLINK2 and bigsnpr. Results are stratified by genetic ancestry of both the testing individuals and the fine-tuning cohorts. The ratio is defined as the Euclidean distance based on PLINK2 PCs divided by that based on bigsnpr PCs.

| **Testing individuals** | **Fine-tuning cohort** | **Pearson’s correlation** | **mean(ratio)** | **sd(ratio)** |
| --- | --- | --- | --- | --- |
| AFR | AFR | 0.999947 | 123.4032 | 3.027889 |
| AFR | EAS | 0.999902 | 122.7597 | 1.664801 |
| AFR | EUR | 0.999946 | 122.1064 | 6.325977 |
| AFR | SAS | 0.999926 | 122.1526 | 2.446598 |
| EAS | AFR | 0.999947 | 123.4032 | 3.027889 |
| EAS | EAS | 0.999902 | 122.7597 | 1.664801 |
| EAS | EUR | 0.999946 | 122.1064 | 6.325977 |
| EAS | SAS | 0.999926 | 122.1526 | 2.446598 |
| EUR | AFR | 0.999947 | 123.4032 | 3.027889 |
| EUR | EAS | 0.999902 | 122.7597 | 1.664801 |
| EUR | EUR | 0.999946 | 122.1064 | 6.325977 |
| EUR | SAS | 0.999926 | 122.1526 | 2.446598 |
| SAS | AFR | 0.999947 | 123.4032 | 3.027889 |
| SAS | EAS | 0.999902 | 122.7597 | 1.664801 |
| SAS | EUR | 0.999946 | 122.1064 | 6.325977 |
| SAS | SAS | 0.999926 | 122.1526 | 2.446598 |
| AMR | AFR | 0.999947 | 123.4032 | 3.027889 |
| AMR | EAS | 0.999902 | 122.7597 | 1.664801 |
| AMR | EUR | 0.999946 | 122.1064 | 6.325977 |
| AMR | SAS | 0.999926 | 122.1526 | 2.446598 |
| OTH | AFR | 0.999947 | 123.4032 | 3.027889 |
| OTH | EAS | 0.999902 | 122.7597 | 1.664801 |
| OTH | EUR | 0.999946 | 122.1064 | 6.325977 |
| OTH | SAS | 0.999926 | 122.1526 | 2.446598 |

Since eventually DiscoDivas used the ratio of Euclidean distance between testing individual and different fine-tuning datasets to calculate the interpolation coefficients, the difference between the interpolate coefficients based on the two sets of Euclidean distance was therefore negligible.

#### Inspection of principal component of ancestry profile among individuals classified as admixed

To assess whether the PRS performance in the OTH samples fairly represents the admixed population—and to rule out the possibility that this performance is primarily driven by a high proportion of European genetic ancestry—we compare the OTH samples with the European (EUR) samples across the three biobanks used for evaluation.

We plotted the heatmaps and density plots of the top 6 PC of individual assigned as “Other” (OTH), American (AMR) and European (EUR) in the three cohorts UKBB, MGBB, and AoU. As shown in the figure, the spatial distribution of densities in OTH more closely resembles that of AMR than EUR. In the UKBB and AoU dataset, we observed that OTH exhibits a density peak near the region covered by UKBB EUR, although this peak was largely separated with the EUR distribution (supplementary figure 13, 14, and 15).

We further tested if the hotspot near EUR area would systematically cause the PRS performance bias. We compared the PRS performance of the quantitative traits only using the UKBB OTH samples outside the host spot area, which is approximately 52% of the overall OTH samples, with the PRS result using all the OTH samples. We observed that in supplementary figure 16, the datapoints were distributed evenly on both sides of the reference line y=x without any obvious enrichment on one side. We therefore conclude that despite the possibility that some of the OTH individuals might have EUR ancestries, the overall distribution of OTH spread across a wide range that are different from EUR and the hotspot near the EUR area in the UKBB OTH sample were unlikely cause obvious bias of the PRS performances.
