## Supplementary Figures with legend for "DiscoDivas: Leveraging genetic ancestry continuum information to interpolate PRS for admixed populations"

Supplementary Figure 1: Comparison of PRS R^2^ of conventional PRS method and DiscoDivas when the fine-tuning dataset for the conventional method is of the matched ancestry with the testing dataset. The four subplots correspond to the four simulated scenarios of varying discovery GWAS sample sizes and causal SNP effect sizes as shown in the 4 panels in Figure 2. Within each subplot, each panel shows the performance of the two methods in each testing sample; the color of the datapoints shows the number of causal SNPs simulated on a base-10 logarithmic scale.

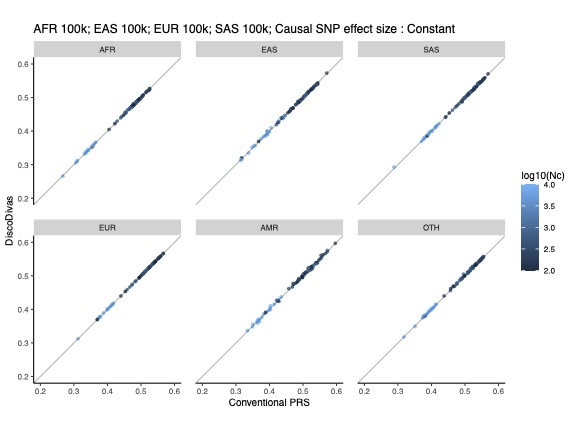

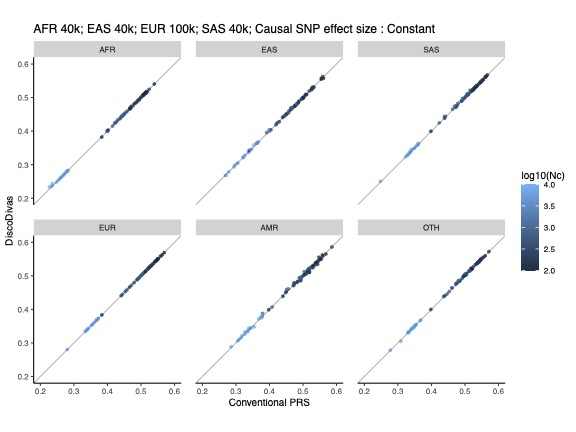

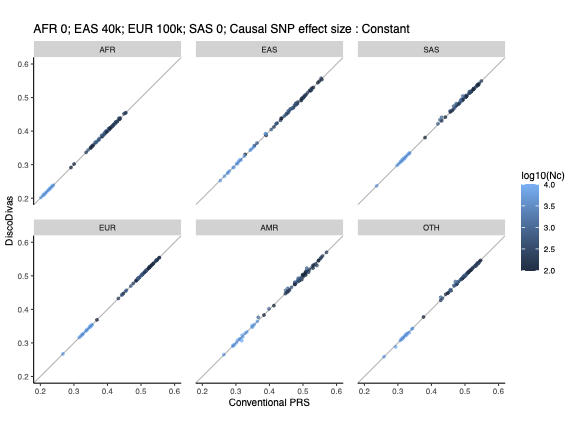

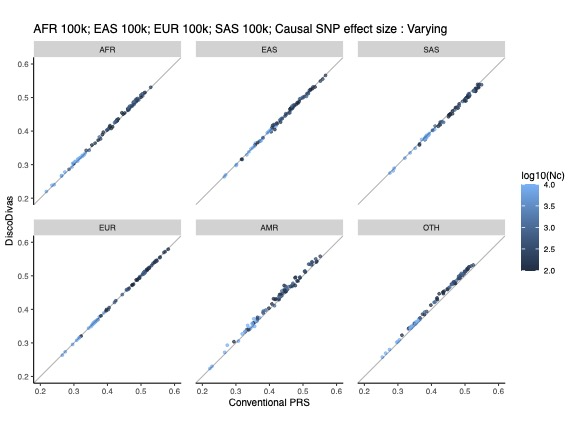

Supplementary Figure 2: The relative increase PRS R^2^ of DiscoDivas over the conventional PRS method. The four subplots correspond to the four simulated scenarios of different discovery GWAS sample sizes and causal SNP effect sizes shown in the 4 panels in Figure 2. Within each subplot, each panel shows the performance of the two methods for each combination of fine-tuning sample for the conventional PRS method and the testing sample; the horizontal bar shows the mean value of the relative increase. The color of the horizontal bar reflects mean value of relative increase and p-value of the paired t-test of DiscoDivas PRS R^2^ and conventional PRS R^2^, with cyan indicating mean increase>0 and p-value<0.0005, dark blue indicating mean increase>0 and p-value<0.05, dark red indicating mean increase<0 and p-value<0.05, bright red indicating mean increase<0 and p-value<0.0005, and grey indicating p-value>0.05

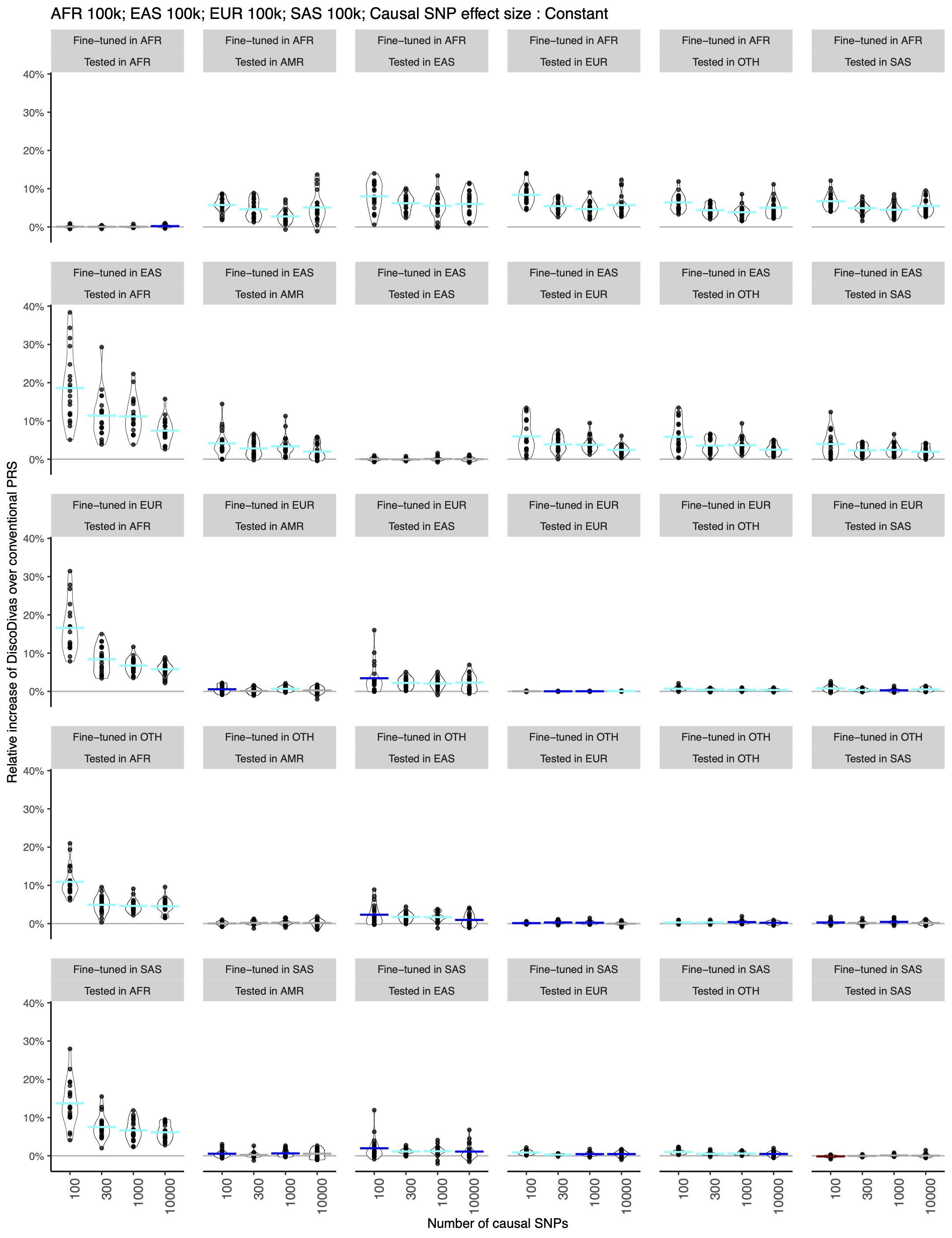

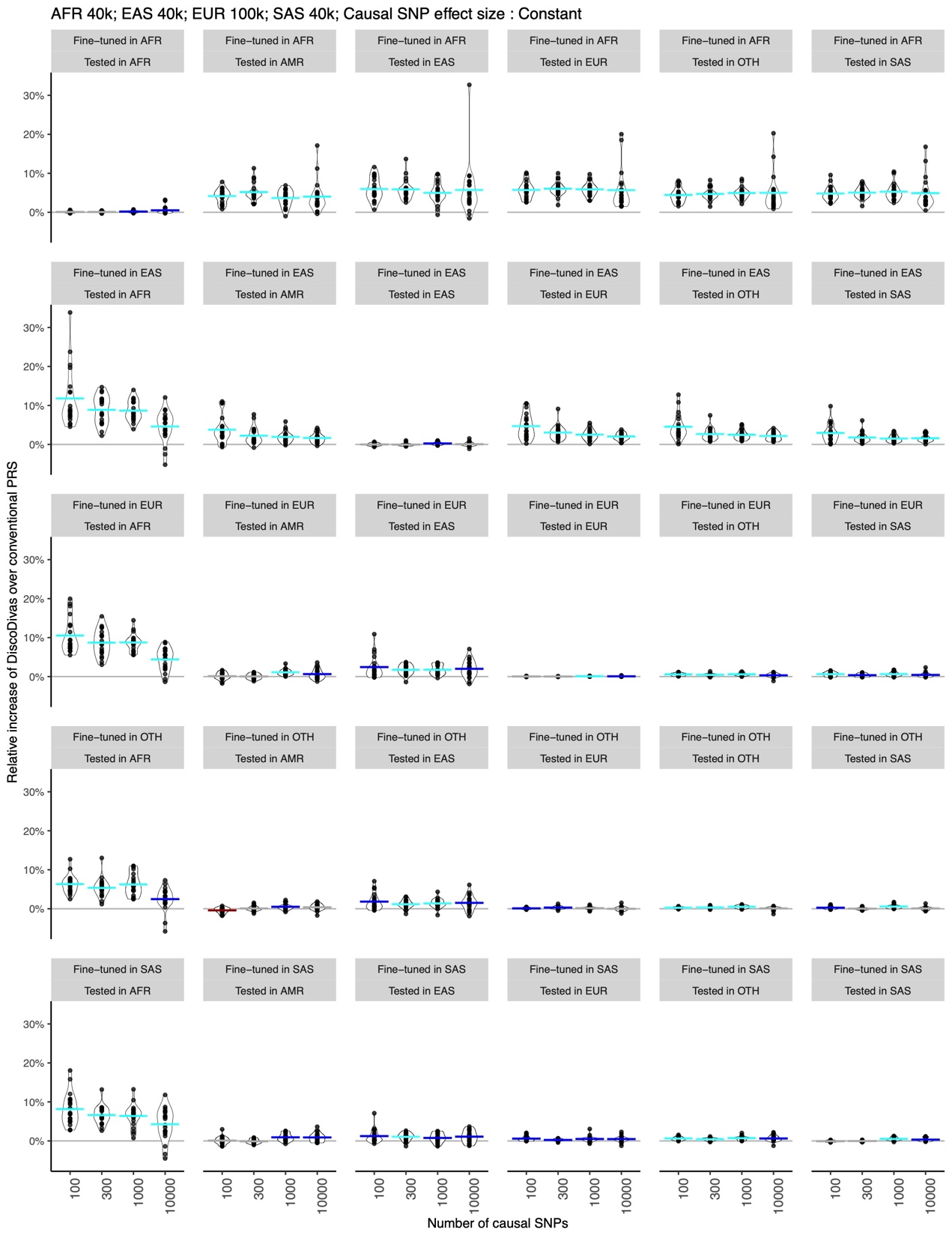

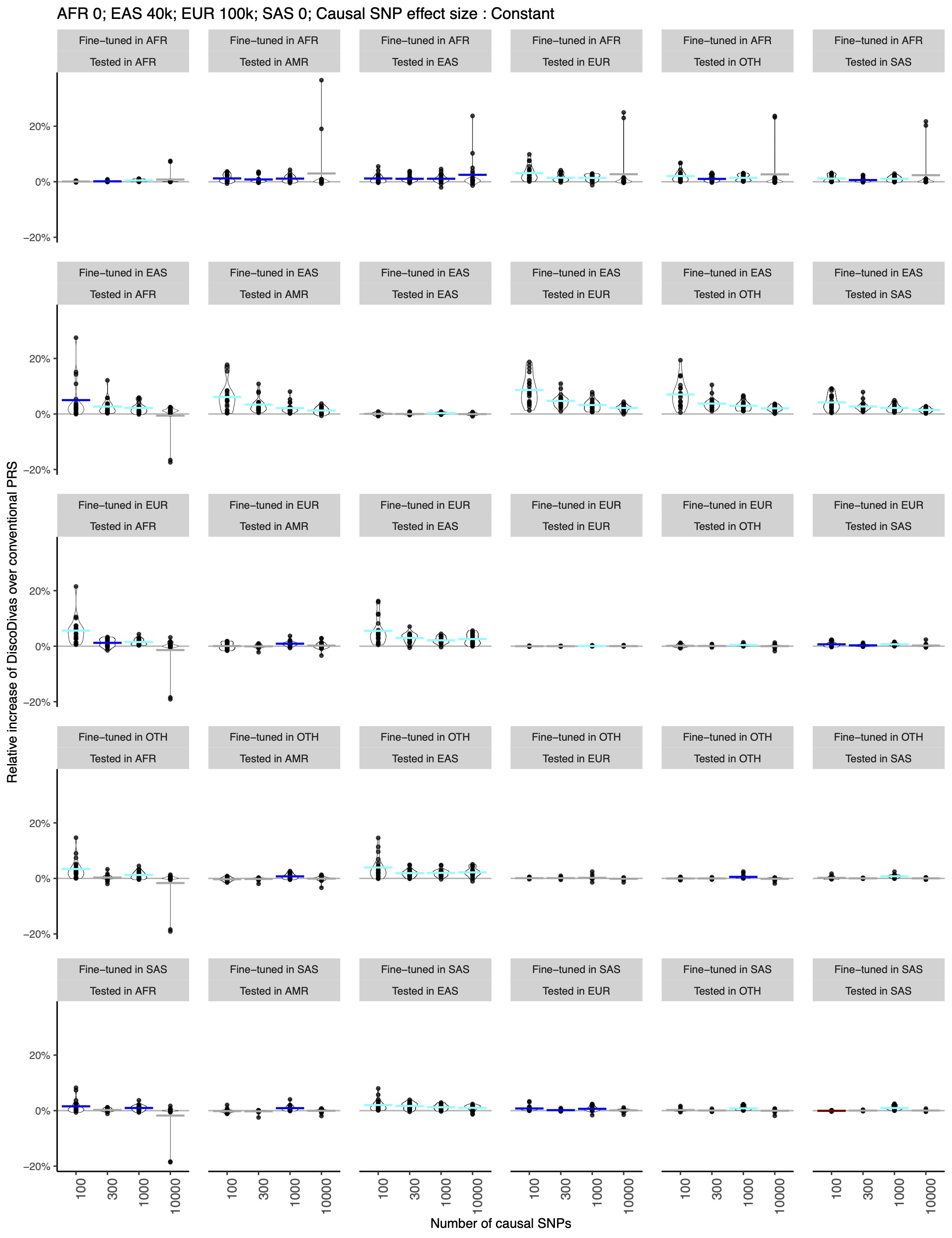

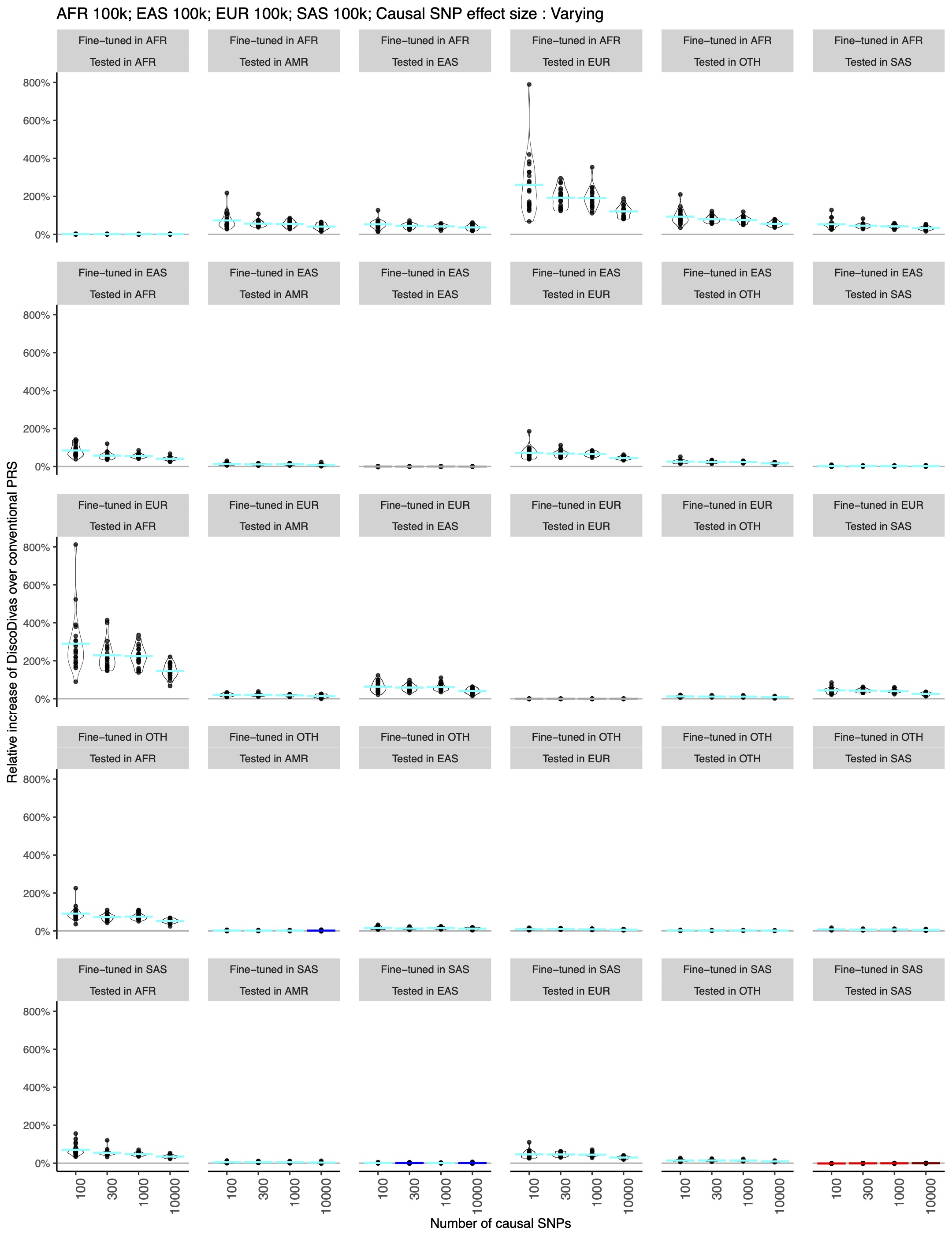

Supplementary Figure 3: Comparison of top 8 PCs of the single-ancestry individuals from the UK Biobank used as fine-tuning sample for DiscoDivas and the 1000 Genomes reference panel. The top row displays the PCs of the 1000 Genomes reference, while the bottom row shows the PCs of single-ancestry individuals from the UK Biobank.

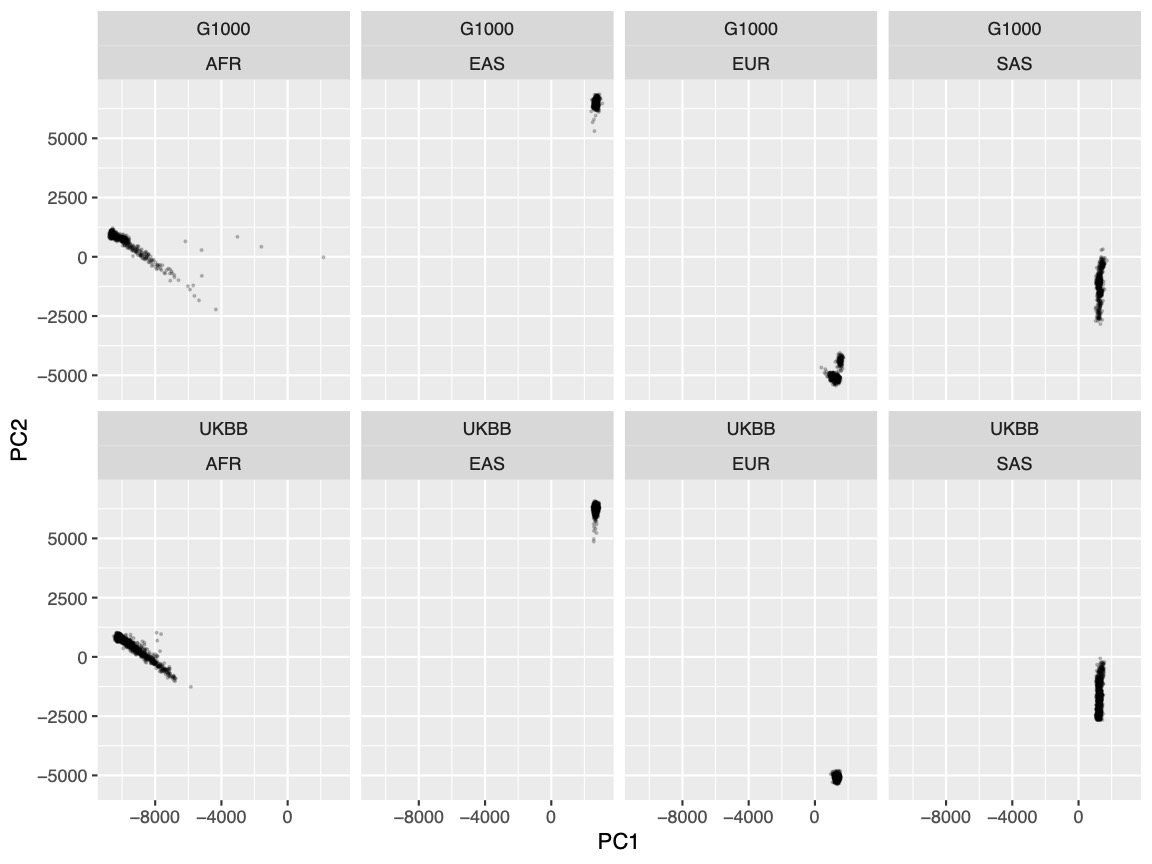

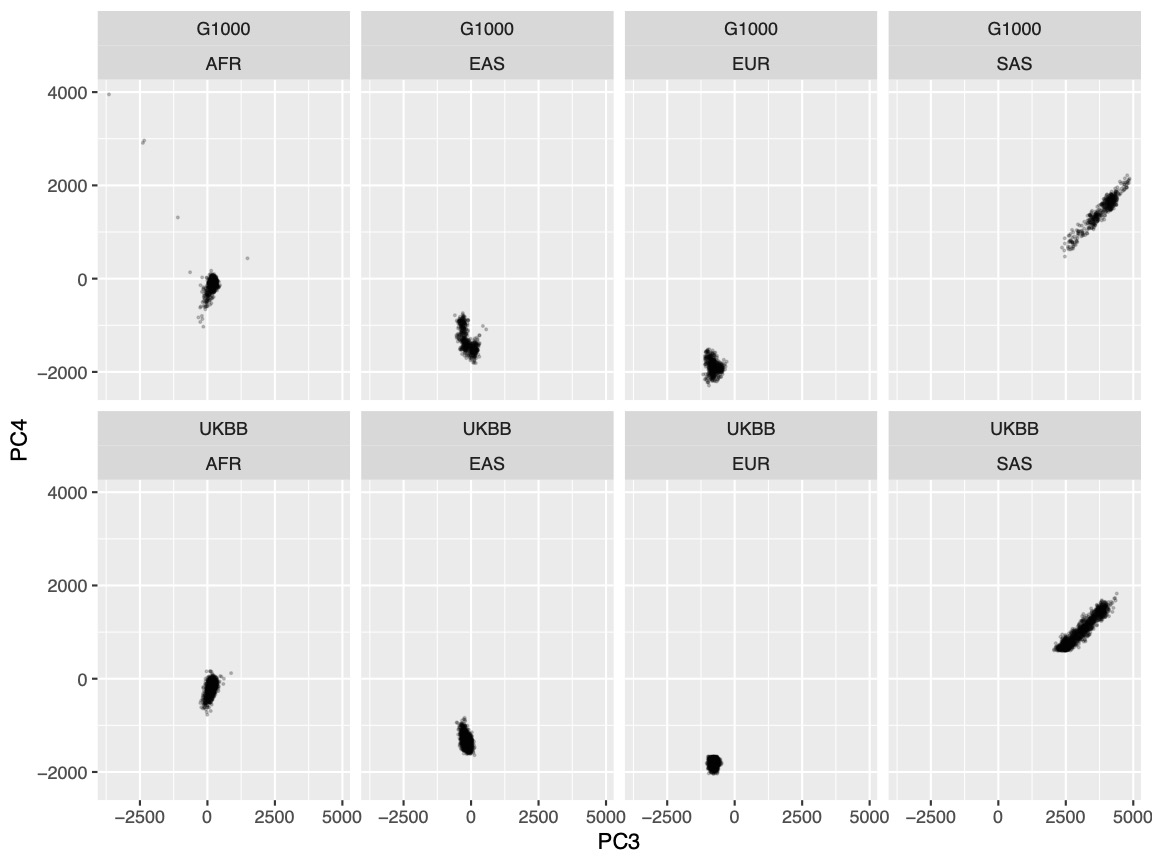

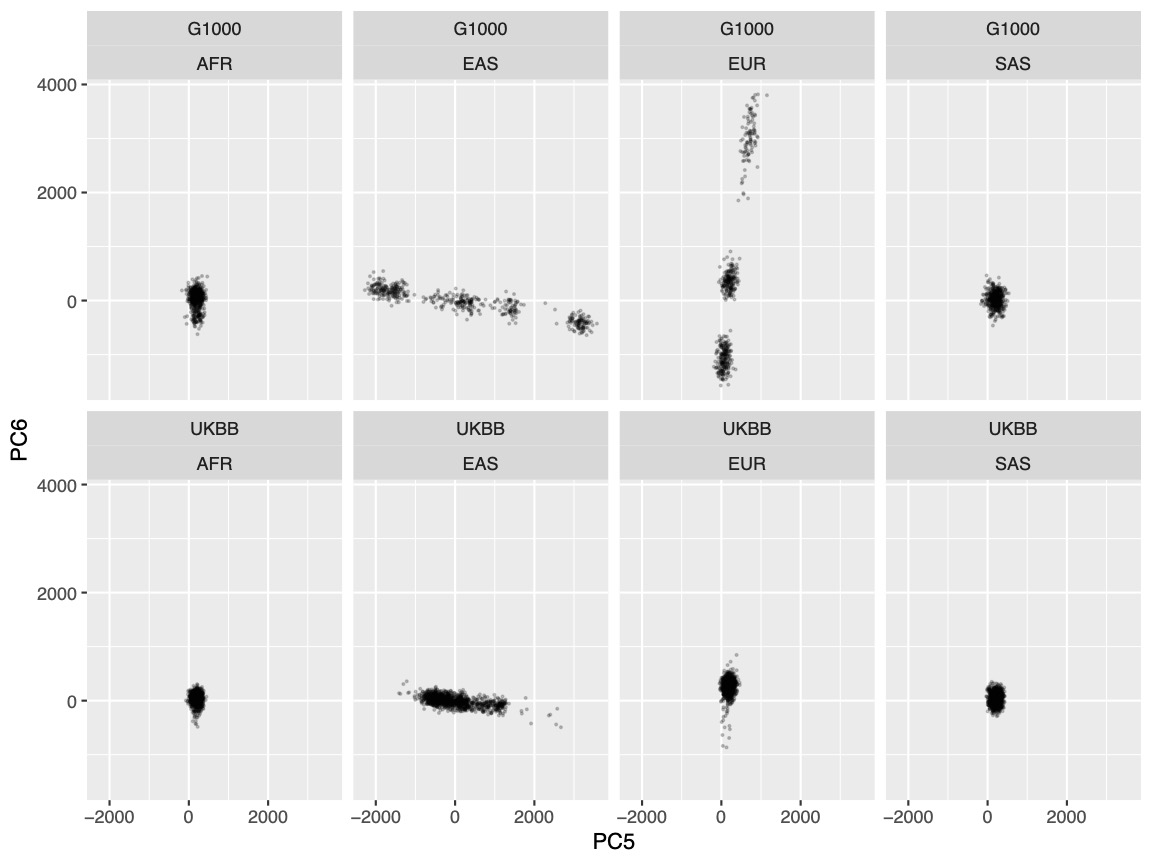

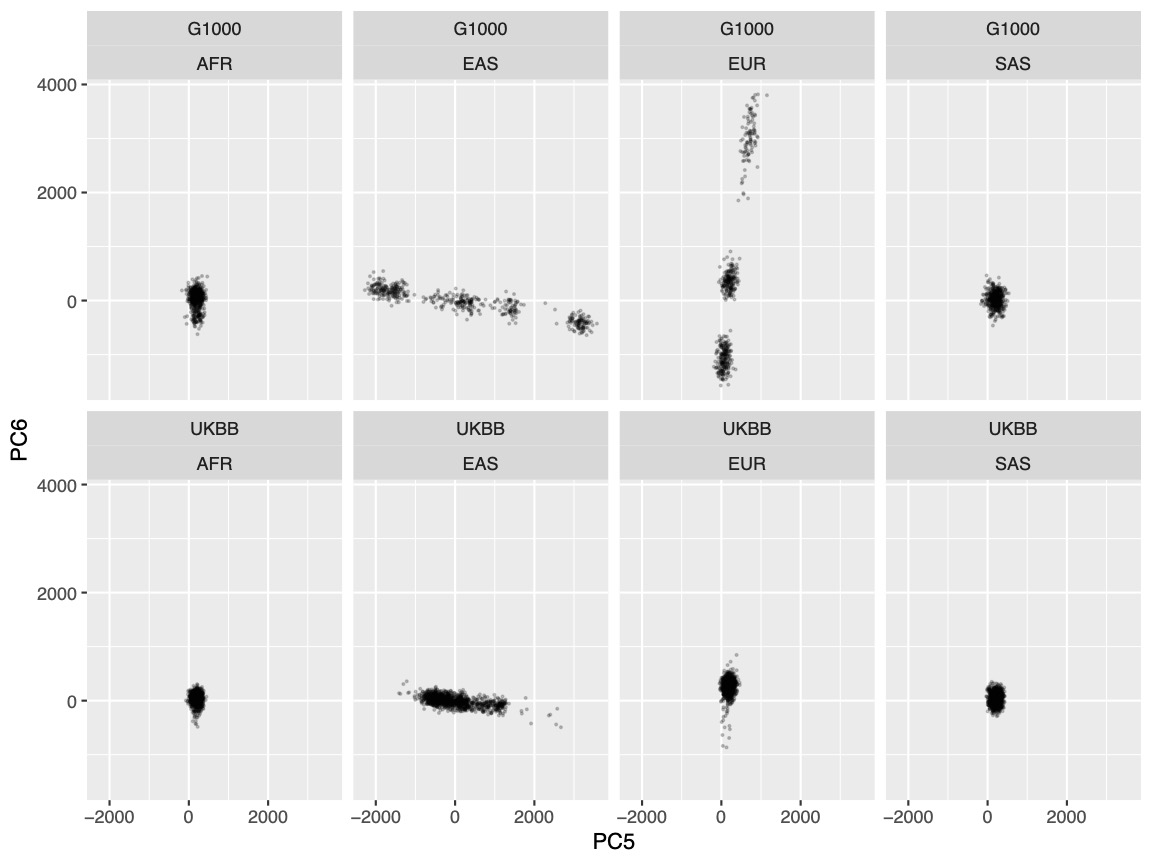

Supplementary Figure 4: The comparison of PCA distance between the testing samples and the median point of fine-tuning, using either the actual PC median point from UKBB fine-tuning samples or an approximated median point based on the 1000 Genomes reference. Each column of the panels shows the ancestry of testing individuals (AFR, EAS, EUR, SAS, AMR and OTH), and each row of the panels shows the distance to the fine-tuning sample $pop$, $D_{pop}$. The range of the distance to median point of the fine-tuning samples is shown along the lower edge of the panel, with the blue color indicating the range of distance based on actual UKBB fine-tuning sample and the red color indicating the range of distance based on 1000 Genomes approximation. $R^{2}$(rsq) of the two sets of calculated distance is shown in each panel.

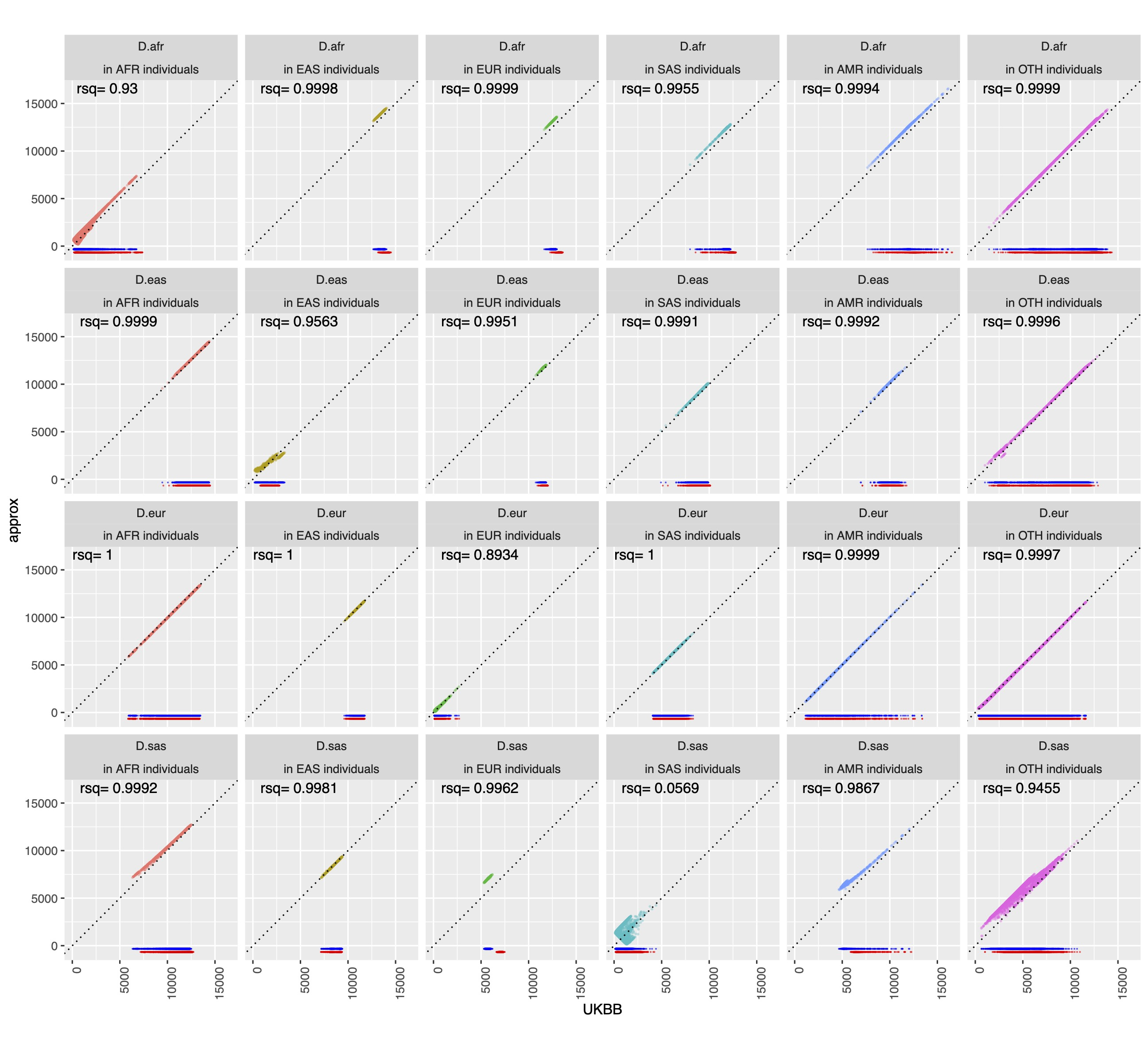

Supplementary Figure 5: The comparison of the interpolation coefficient of the input PRS used in DiscoDivas based on the PC of UKBB fine-tuning samples versus based on the approximated PCA of the 1000 Genomes reference. Each column of the panels shows the ancestry of testing individuals (AFR, EAS, EUR, SAS, AMR and OTH), and each row of the panels shows the interpolation coefficient $w_{pop}$ of the input PRS based on the four UKBB single-ancestry fine-tuning datasets (AFR, EAS, EUR, and SAS). The range of combination coefficients is shown along the lower edge of the panel, with the blue color indicating the range of combination coefficients based on actual UKBB fine-tuning sample and the red color indicating the range of combination coefficients based on 1000 Genomes approximation. $R^{2}$ (rsq) of the two sets of combination coefficients is shown in each panel.

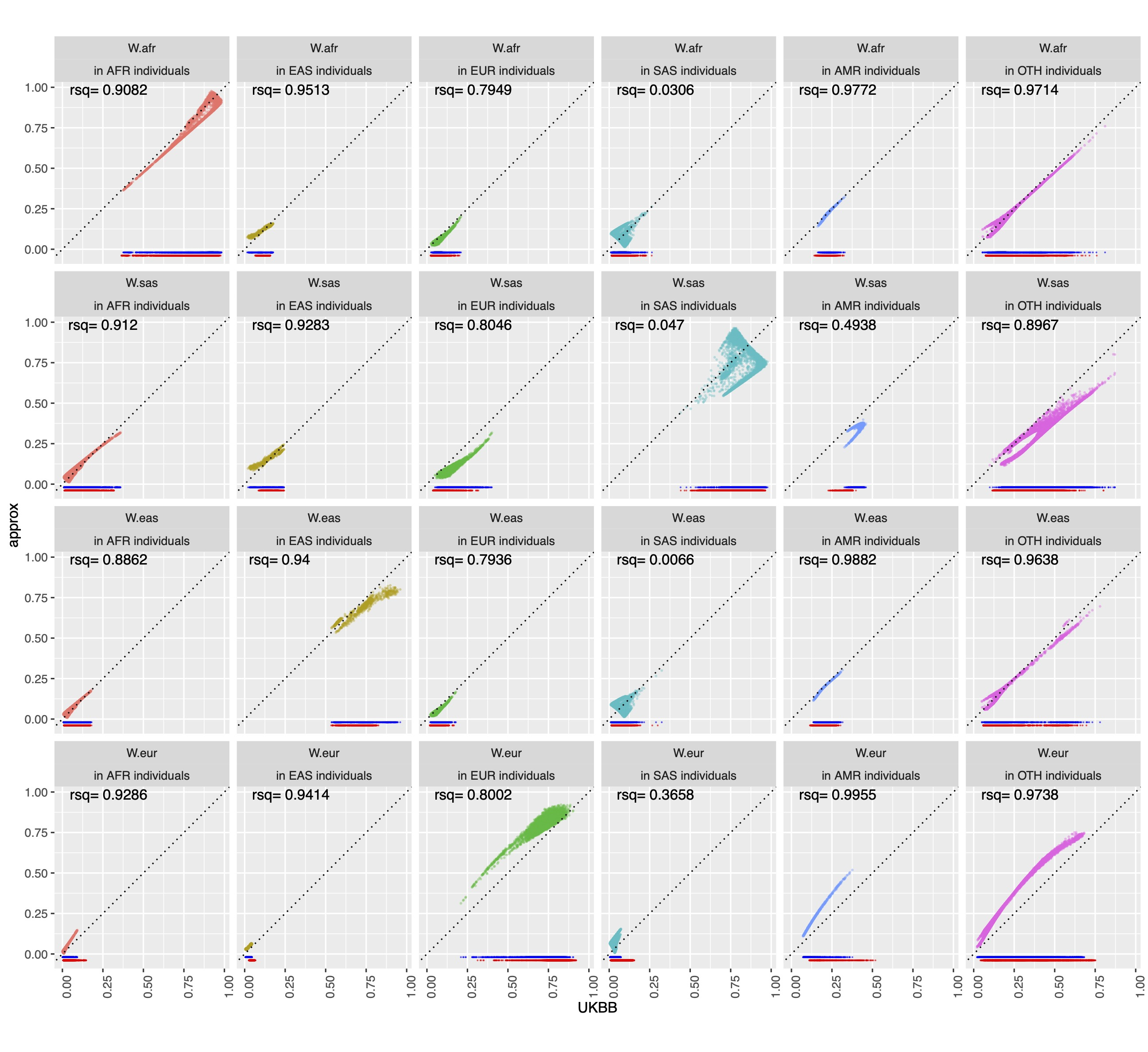

Supplementary Figure 6: The comparison of DiscoDivas PRS R^2^ when using the PCA value of UKBB fine-tuning samples and the approximated PCA value of using the 1000 Genomes reference. Each column of the panels shows the ancestry of testing individuals, and each row of the panels and the color of datapoints show the simulated number of causal SNPs. The four subplots correspond to the four simulated scenarios of different discovery GWAS sample sizes and causal SNP effect sizes shown in the 4 panels in Figure 2.

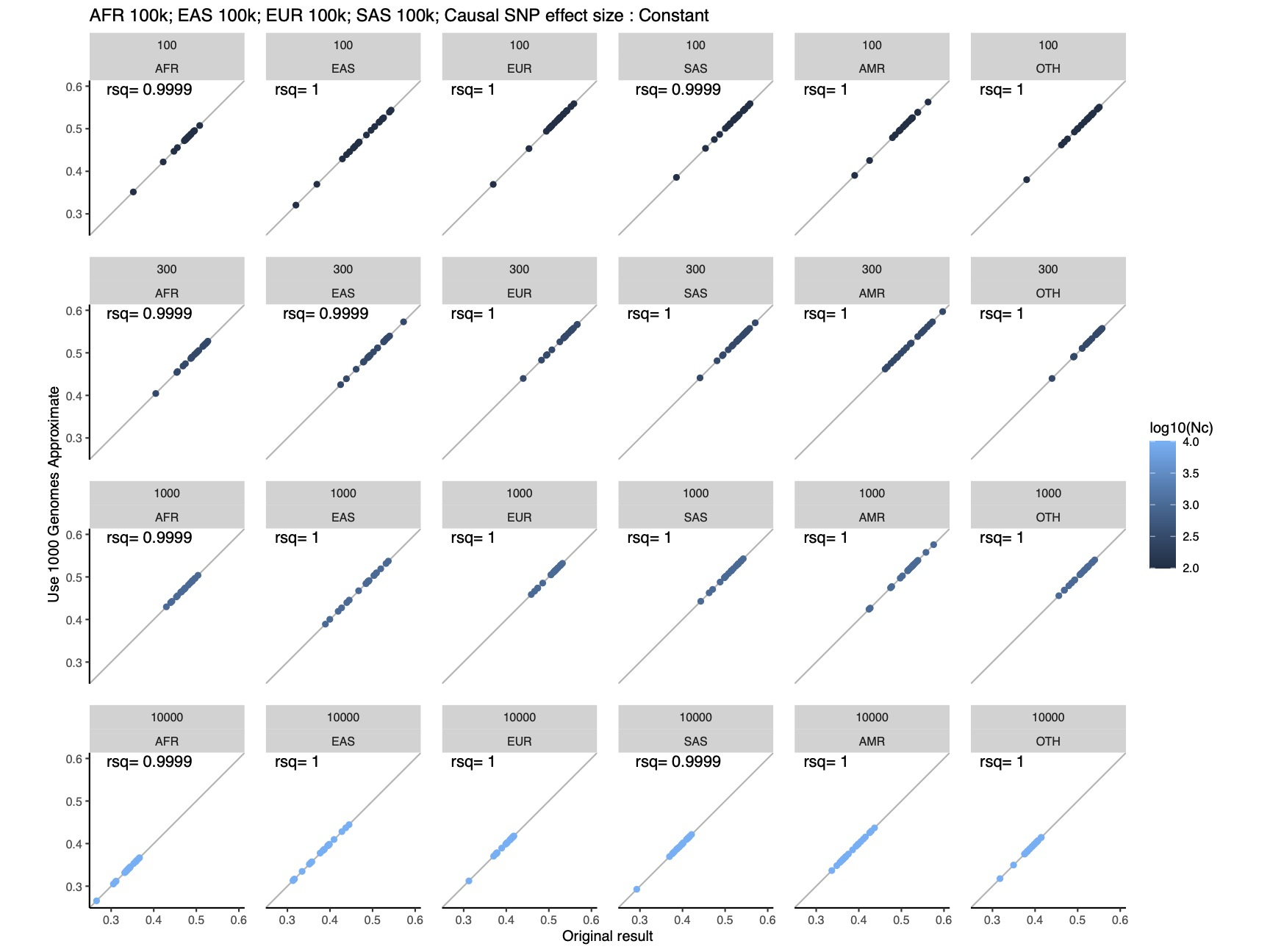

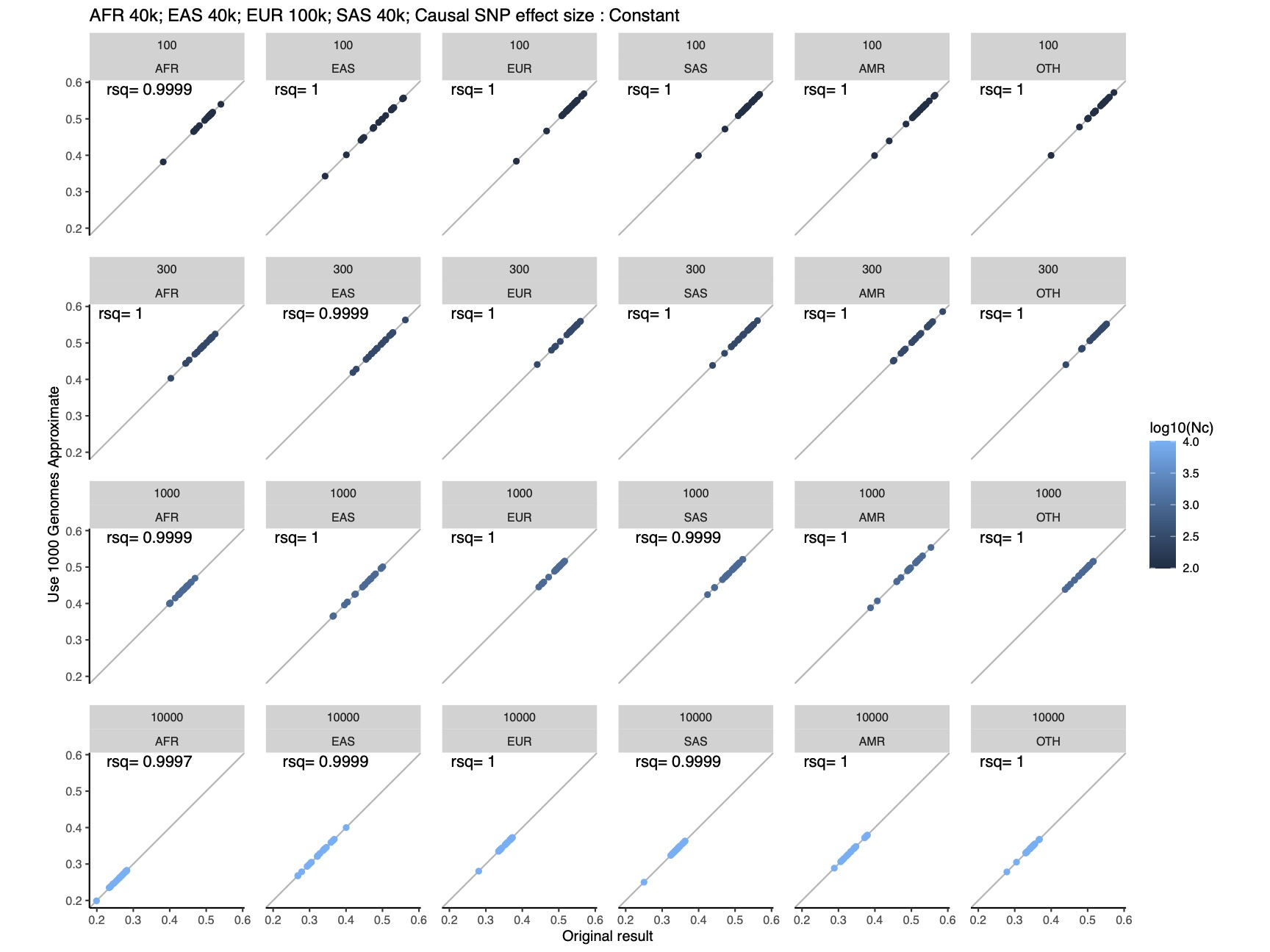

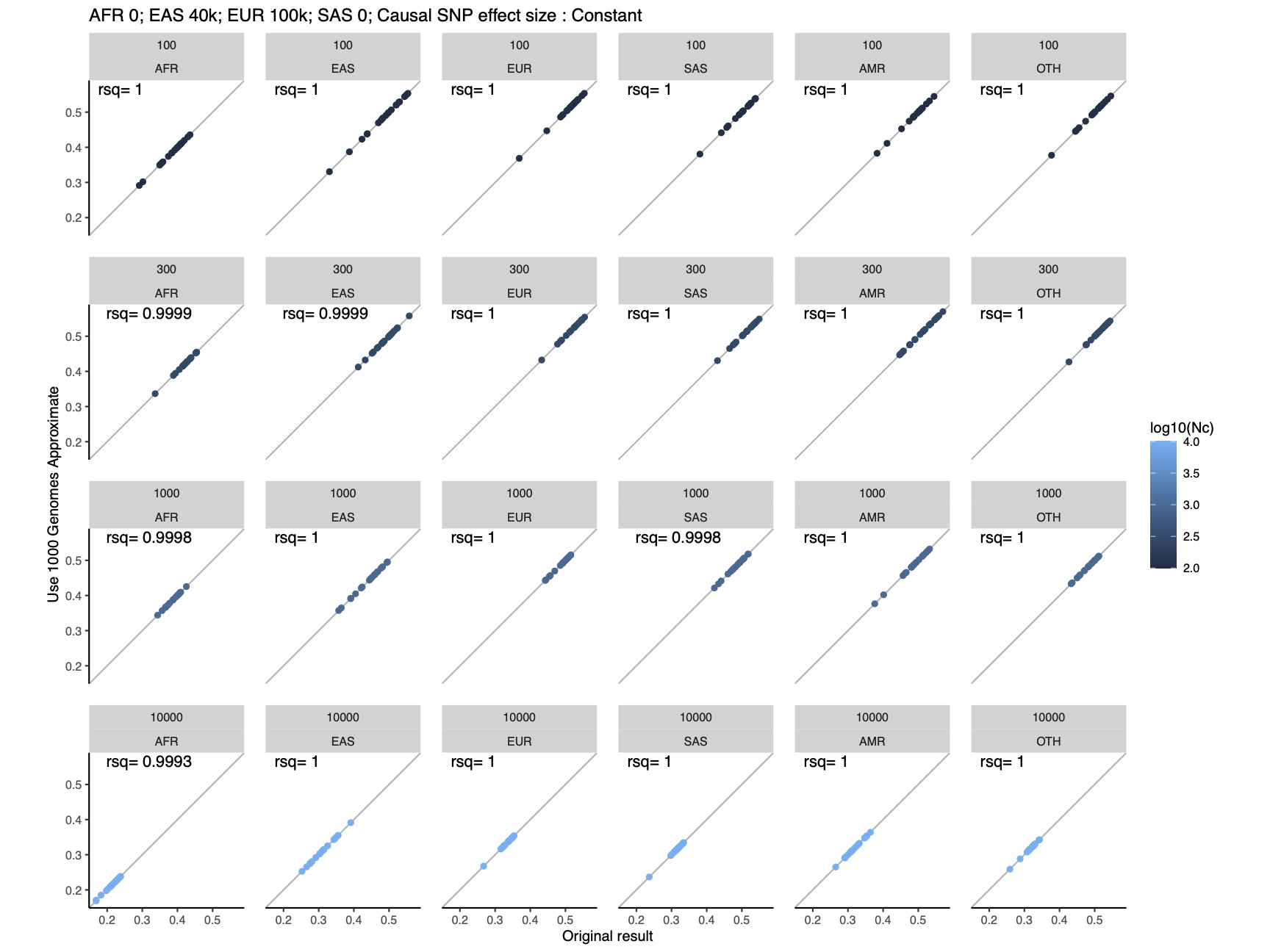

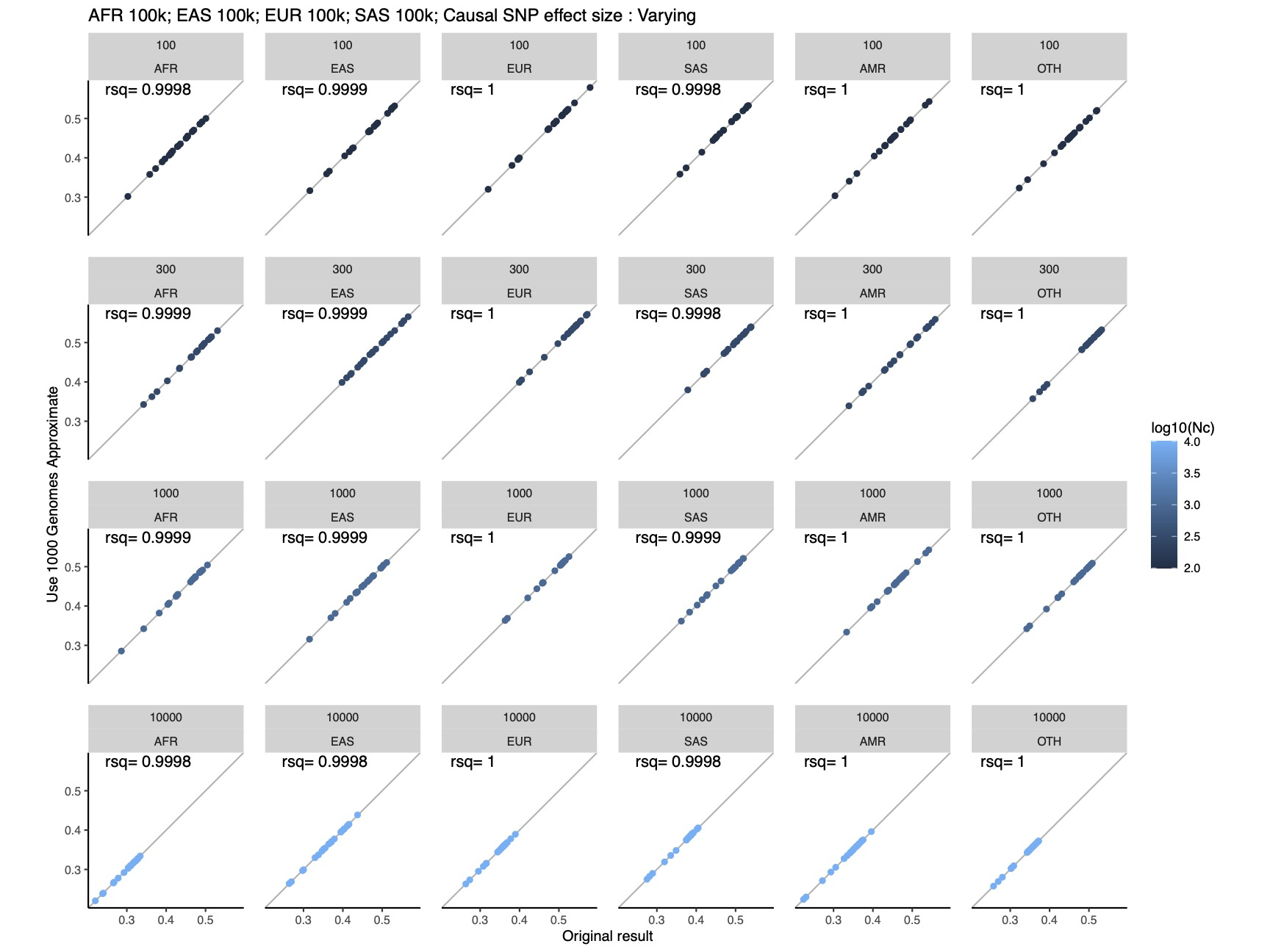

Supplementary Figure 7 Comparison of PRS R^2^ of using UKBB-based fine-tuning samples and the purely simulated fine-tuning samples. The upper subplot shows the results of the conventional PRS method and the lower shows the result of DiscoDivas. Within each subplot, the column of panels shows the ancestry of testing samples and the row of the panels and the color of datapoints show the simulated number of causal SNPs. The x-axis shows the PRS R^2^ using UKBB-based fine-tuning samples and the y-axis shows the PRS R^2^ using simulated fine-tuning samples.

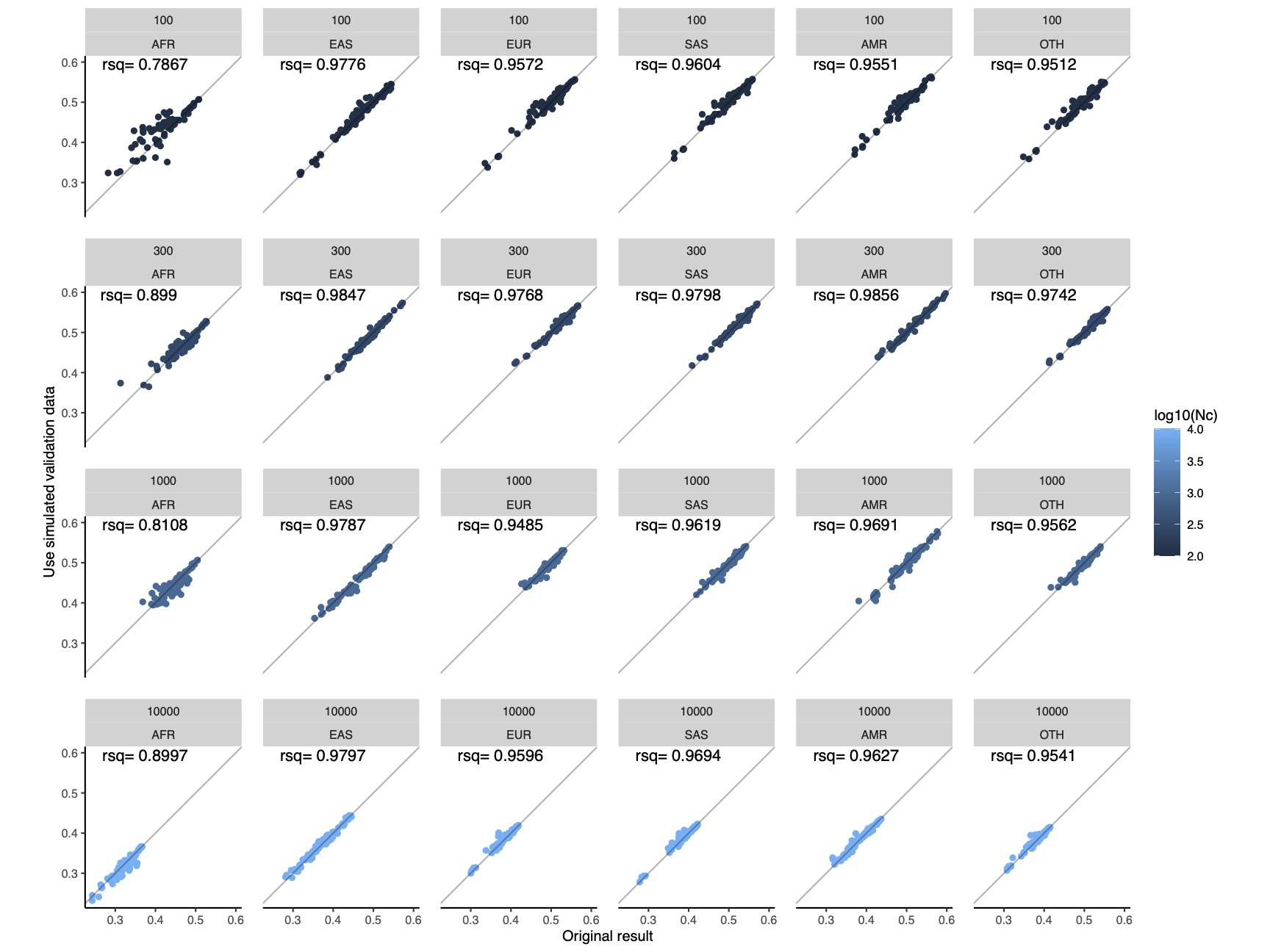

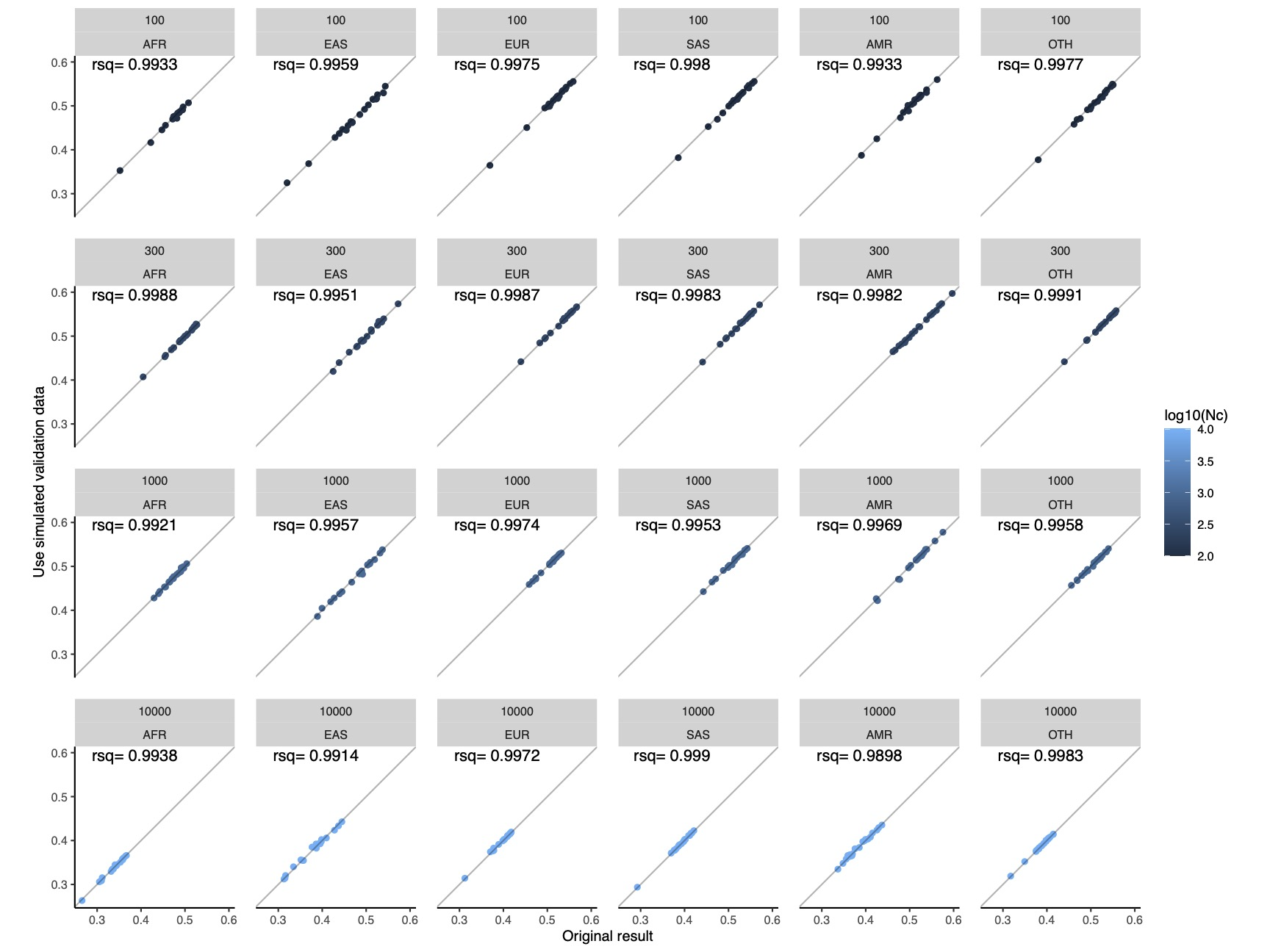

Supplementary Figure 8: The relative increase PRS R^2^ of DiscoDivas over the conventional PRS method when using the purely simulated fine-tuning data. Each panel shows the performance of the two methods in each combination of fine-tuning sample for the conventional PRS method and the testing sample; the horizontal bars show the mean value of the relative increase. The color of the horizontal bar indicates mean value of relative increase and p-value of the paired t-test of DiscoDivas PRS R^2^ and conventional PRS R^2^, with cyan indicating mean increase>0 and p-value<0.0005, dark blue indicating mean increase>0 and p-value<0.05, dark red indicating mean increase<0 and p-value<0.05, bright red indicating mean increase<0 and p-value<0.05, and grey indicating p-value>0.05

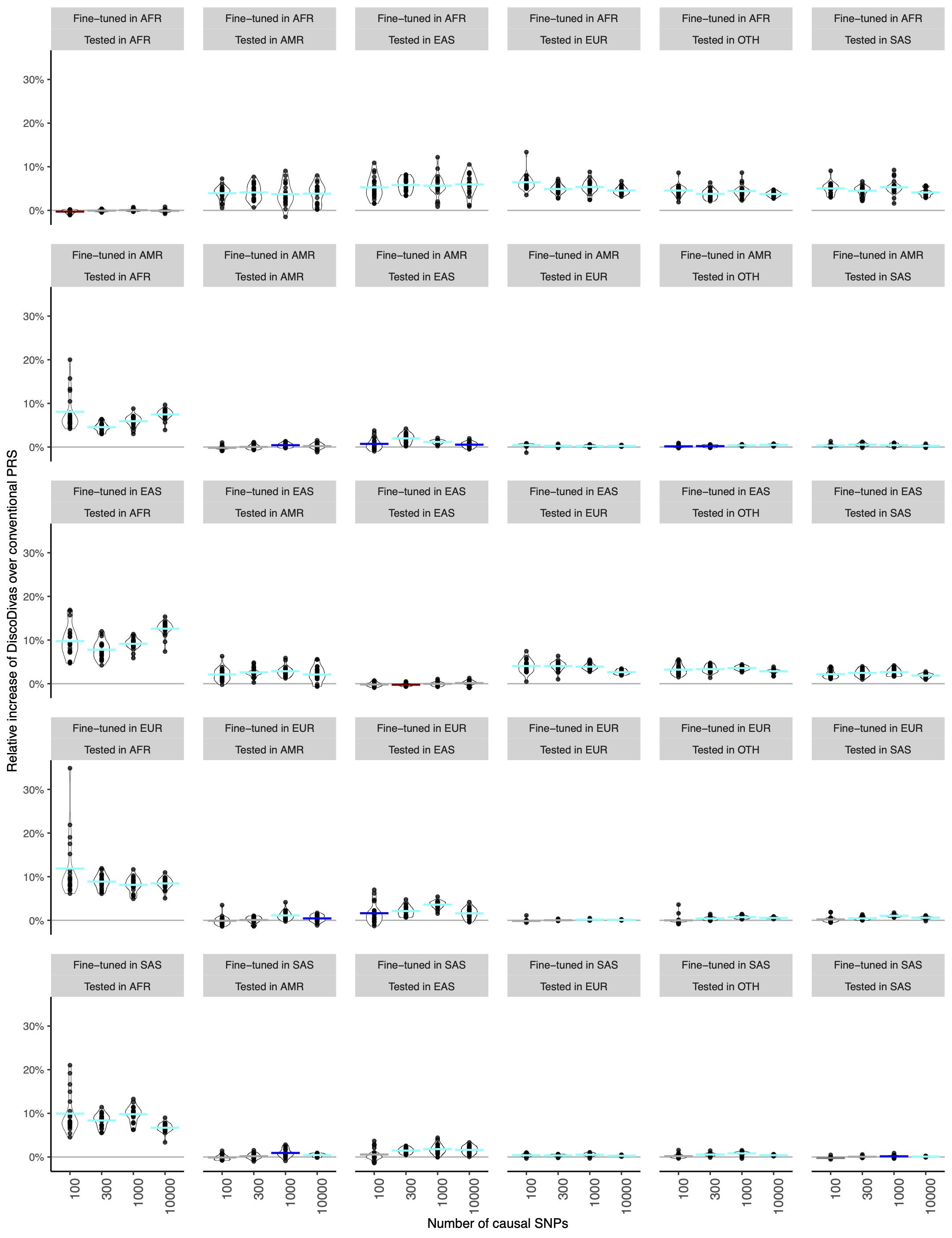

Supplementary Figure 9: The PRS distance between individuals in testing samples to the median point of the fine-tuning samples when including different numbers of PCA in the distance calculation. Each panel shows the ancestry of the testing samples and the color of the line shows the fine-tuning samples to which the distance is calculated. The plot is based on 100 randomly selected individuals from each UKBB testing sample.

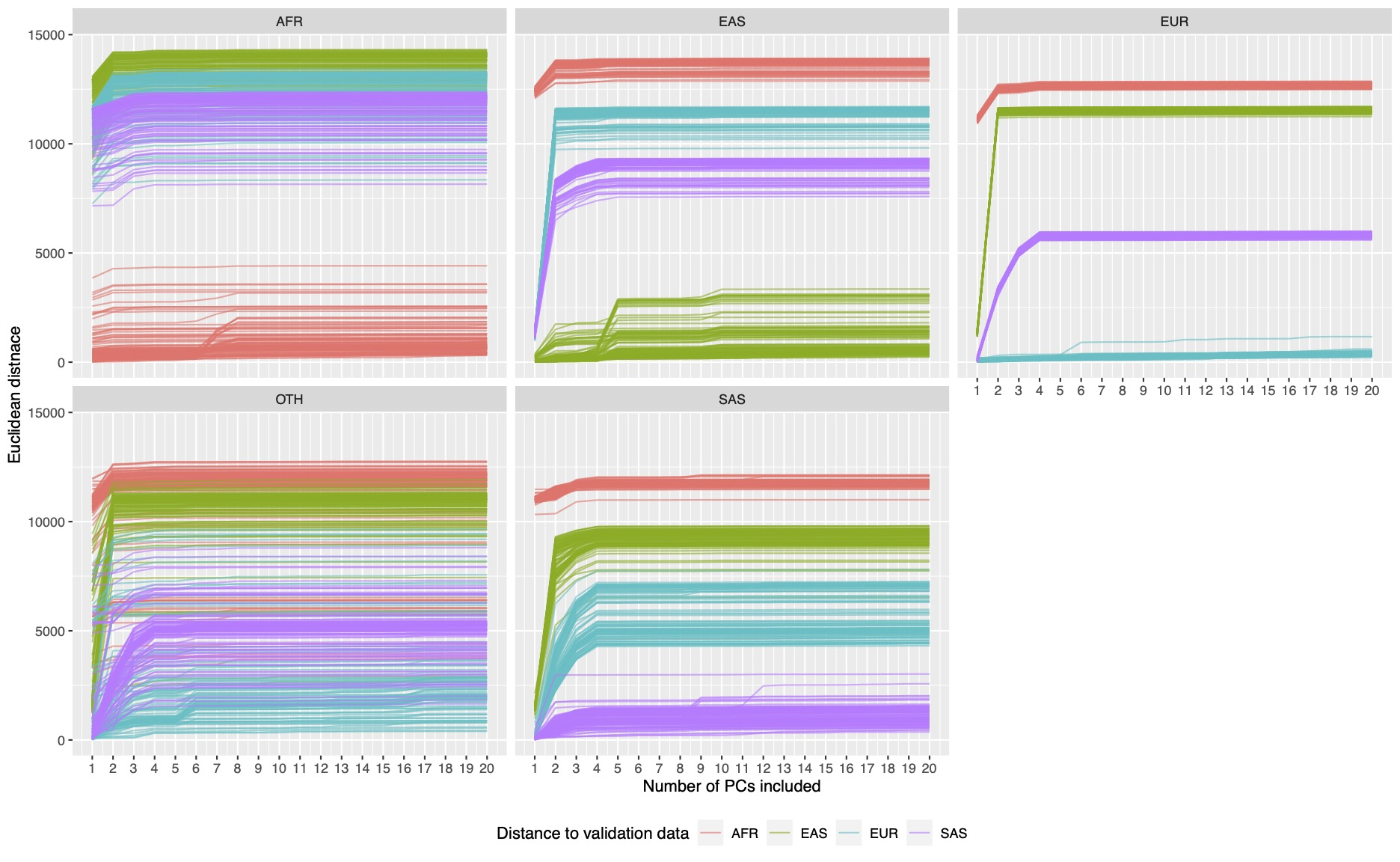

Supplementary Figure 10: The alternative DiscoDivas workflow of using single-population PRS as the input. Each input PRS was based on a single-population GWAS and fine-tuned using the matched population data. Note that in the scenario where only EUR and EAS discovery GWAS were available, DiscoDivas was based on only EUR and EAS PRS.

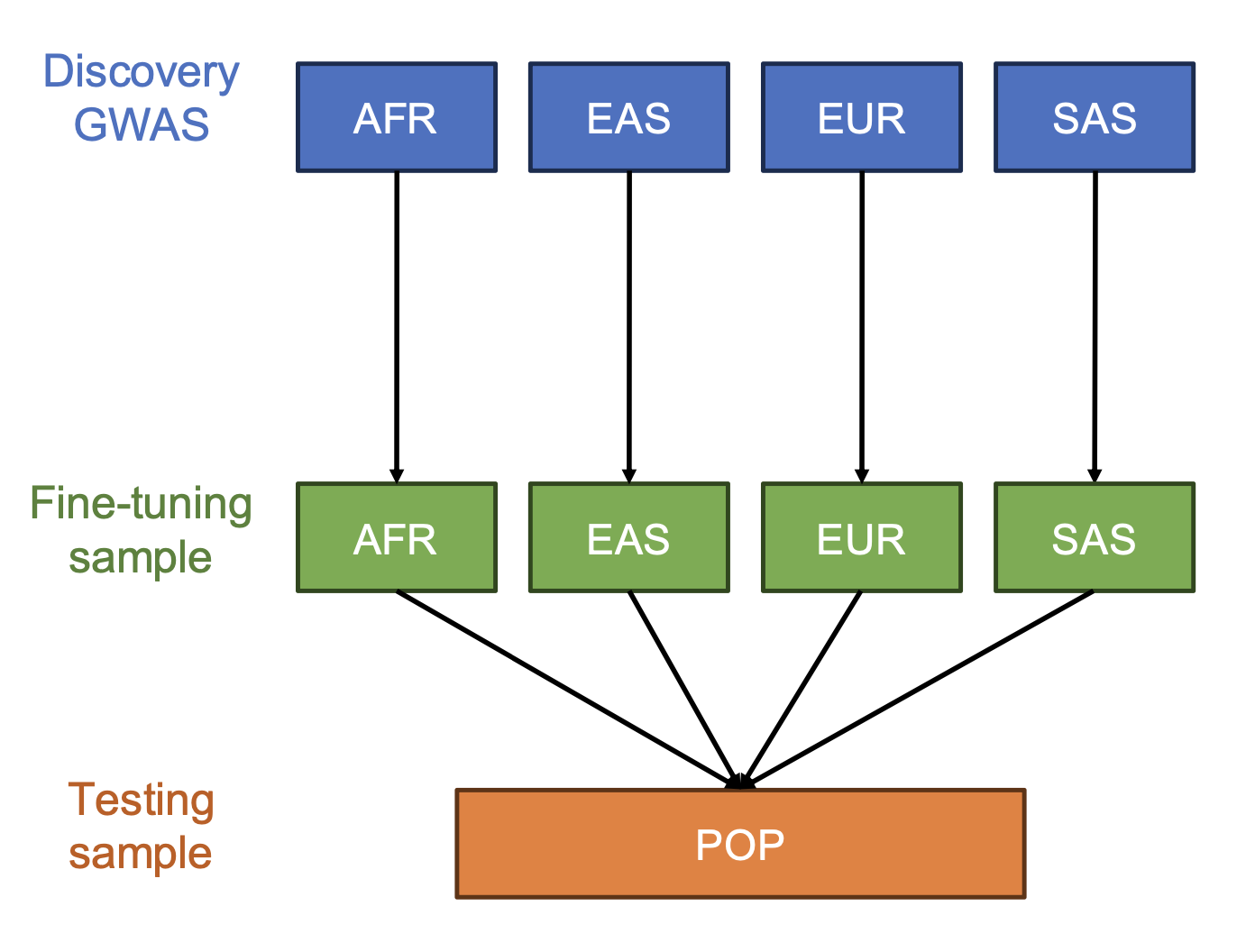

Supplementary Figure 11: The PRS accuracy of different approaches tested in simulated data based on the genotype of UKBB admixed samples. The plot is based on same discovery GWAS as used in Figure 2 of the main text. The four panels show the input data combination, the subplot in each panel shows the number of simulated casual SNPs, and the y-axis shows the PRS R^2^. The four approaches are the single-population PRS that performed the best in the corresponding fine-tuning data, DiscoDivas PRS based on single-population PRS, multi-population PRS fine-tuned in the matched population, and DiscoDivas PRS based on multi-population PRS.

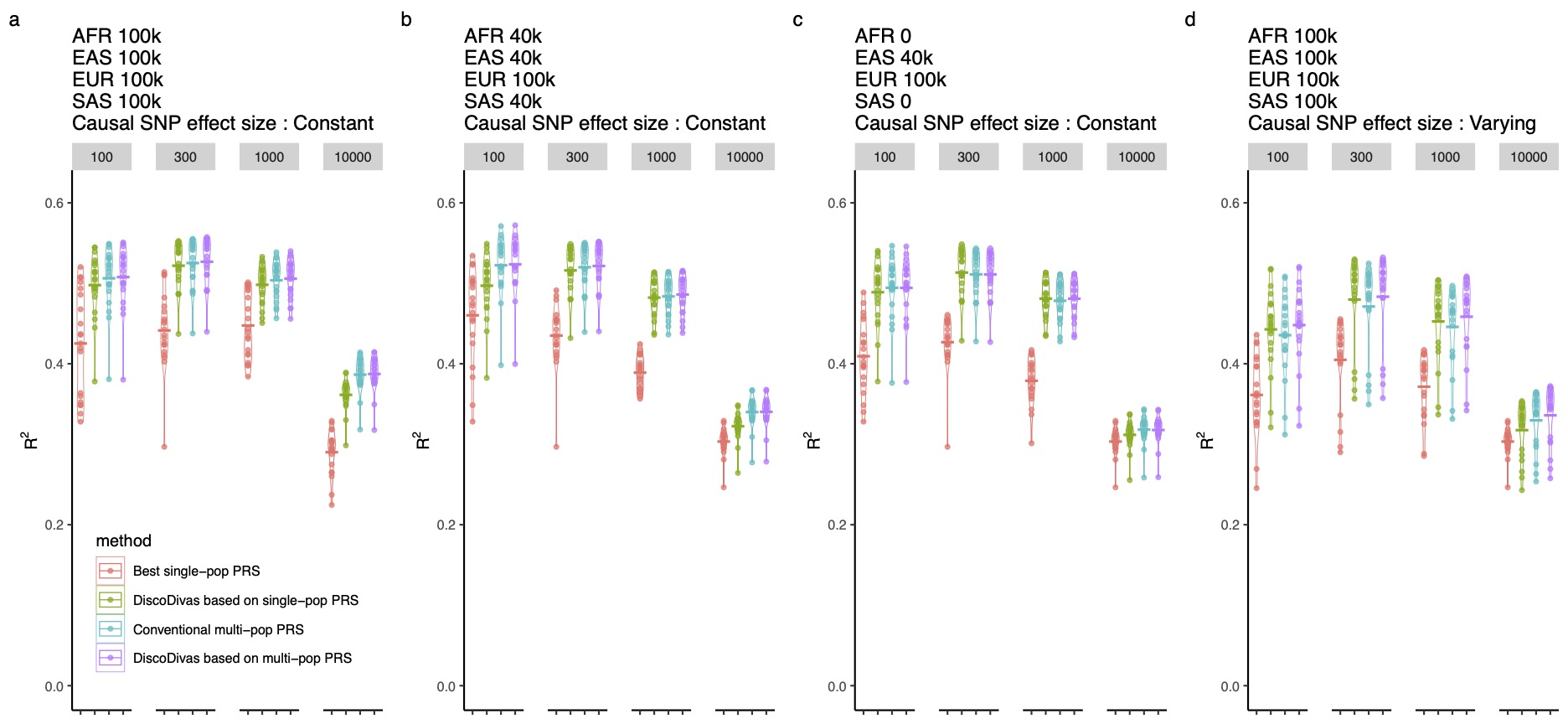

Supplementary Figure 12: Absolute values of Pearson’s correlation coefficients between the top 20 principal components of ancestry computed using PLINK2 and **bigsnpr**.
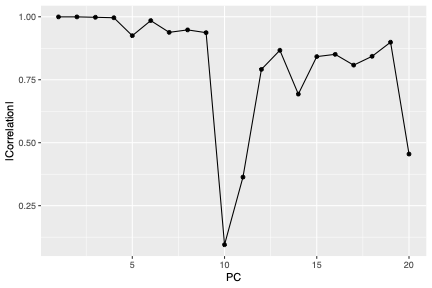

Supplementary figure 13: Density plots showing the distribution of UKBB individuals from EUR, AMR, and OTH ancestry groups in PC space. To enable visual comparability, all AMR and OTH individuals were included, while EUR individuals were down-sampled to one-third the total number of AMR and OTH individuals.

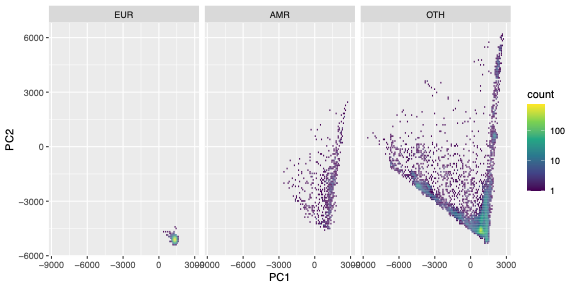

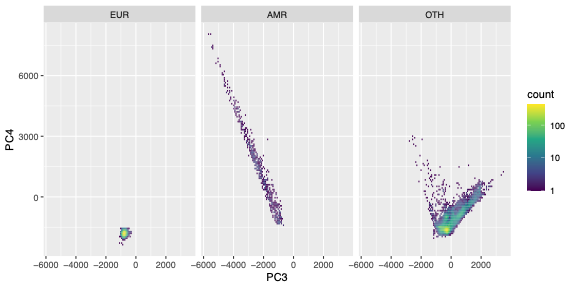

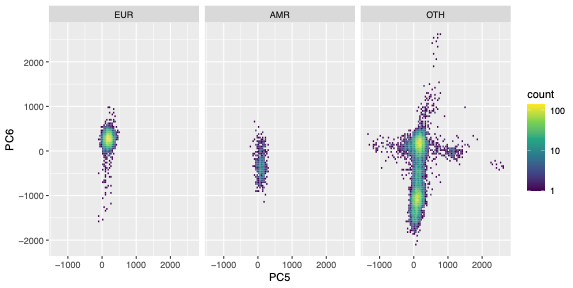

Supplementary figure 14: Density plots showing the distribution of MGBB individuals from EUR and AMR ancestry groups in principal component (PC) space. To enable visual comparability, all AMR individuals were included, while EUR individuals were down-sampled to one-third the total number of AMR individuals.

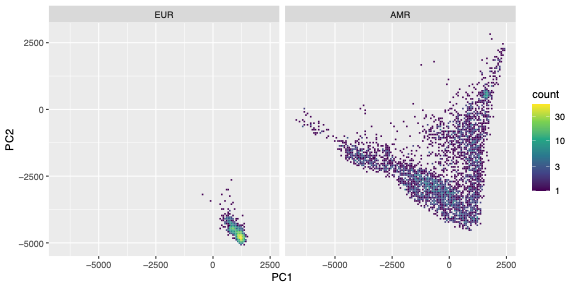

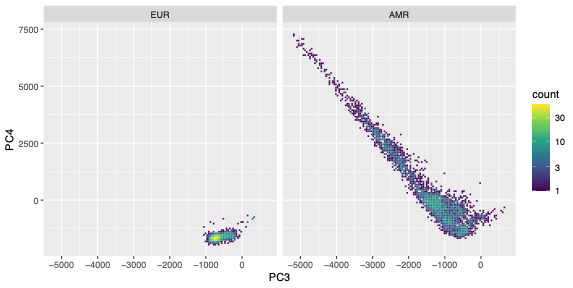

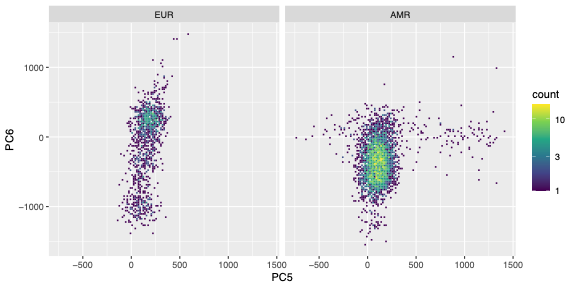

Supplementary figure 15: Density plots showing the distribution of AoU individuals from EUR and AMR ancestry groups in principal component (PC) space. To enable visual comparability, all AMR individuals were included, while EUR individuals were down-sampled to one-third the total number of AMR individuals.

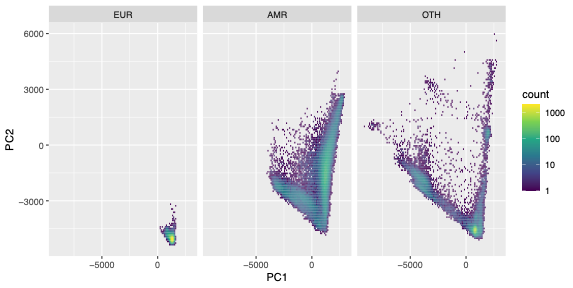

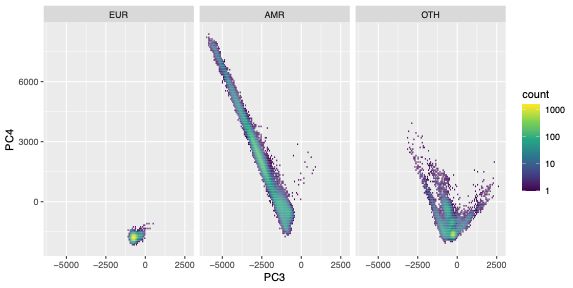

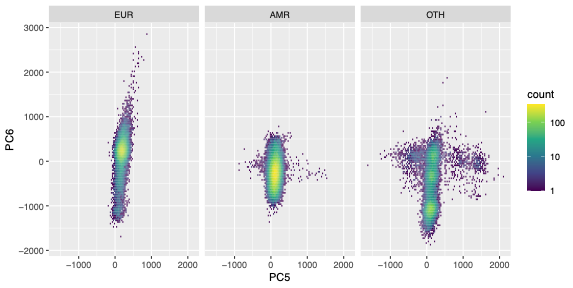

Supplementary Figure 16: Comparison of R^2^ for quantitative trait PRS in all OTH individuals versus OTH individuals outside the EUR-adjacent hotspot in UKBB ancestry PC space.

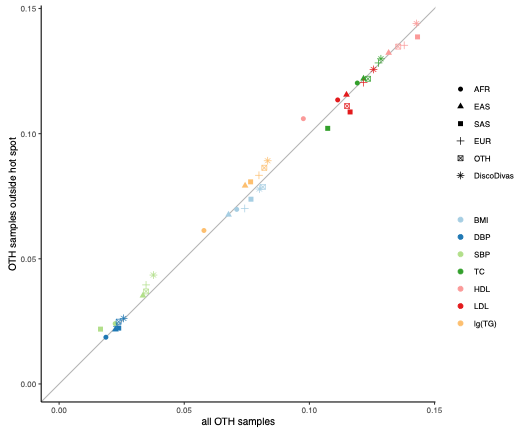

Supplementary figure 17: Relative R2 increase of DiscoDivas over the conventional PRS fine-tuned in one single sample. The panel shows the population in with the PRS was tested and x-axis shows the population in which the PRS was fine-tuned. The horizontal bar shows the mean of relative increase, and the line-type of the bar indicates the p-value of paired t-test of DiscoDivas PRS R2 and conventional PRS R2, with the solid bar being p-value <0.05 and dotted bar being p-value>0.05.

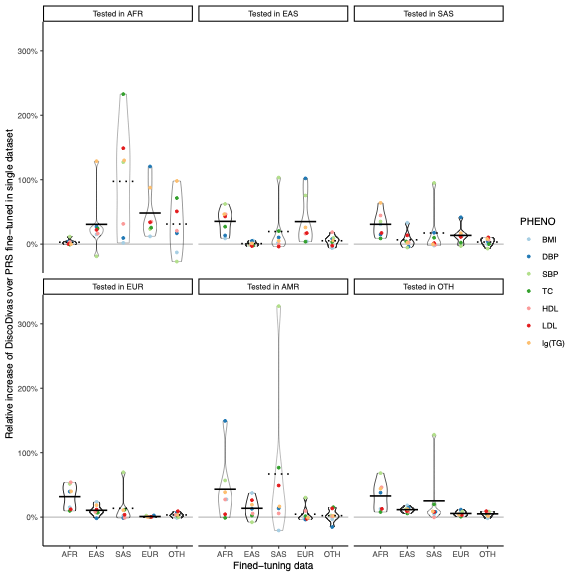
